# Real-world deployment of remote sleep monitoring technologies reveals distinct patterns associated with cognitive decline

**DOI:** 10.1101/2025.08.29.25334735

**Authors:** Nan Fletcher-Lloyd, Nathalia Céspedes Gómez, Alexander Capstick, Antigone Fogel, Marirena Bafaloukou, Mahan Heydari, Alexandra Cairns, Chloe Walsh, Jessica True, CR&T Group, Behnam Shariati, Ramin Nilforooshan, Payam Barnaghi

## Abstract

**BACKGROUND:** Sleep disturbances and altered circadian rhythms are well-documented in both physiological and biological studies of dementia. The exact causal relationship remains unclear. Several other long-term health conditions may also influence sleep patterns. Examining sleep patterns in relation to chronological ageing and the variations and progression of dementia, considering factors such as age, sex, and disease stage, can offer new insights for developing early screening and risk identification measures. Remote sleep monitoring enables routine assessment of cognitive decline symptoms in high-risk groups, aiding early risk identification. Integrating predictive models with routine sleep data holds promise for shaping more proactive approaches to assessing and caring for older adults and people living with dementia (PLWD).

**METHODS:** We developed a machine learning pipeline to estimate Sleep Age Index (SAI) from longitudinal remote sleep monitoring data collected using under-the-mattress sleep sensors and use it to identify dementia risk. The study utilised a dataset of nocturnal activity and physiology data (n=1,672; 18,369 person-samples) collected from individuals in the general population and a cohort of PLWD. Dementia risk scores were stratified into high, medium and low-risk categories to support clinical monitoring and decision-making.

**RESULTS:** Our study indicates that sleep patterns in dementia do not follow typical ageing processes. For individuals with dementia, our pre-trained machine learning model predicted age in a negative direction. Further investigation revealed distinct patterns associated with these predictions, including irregular times to bed and rise, lower variation in deep sleep duration, and elevated breathing and heart rates. While age predictions were largely similar for female and male participants, the patterns observed in the female group were more consistent. Chronological age was predicted from sleep data with a mean absolute error of 5.52 (95% CI: 5.37 - 5.67) on held-out test data. A sensitivity of 75.7% (95% CI: 71.4% - 79.9%) and specificity of 74.7% (95% CI: 69.2% - 80.0%) was achieved post-stratification on unseen data in dementia versus control. To evaluate the real-world clinical applicability of the model, we conducted a pilot study in a population with a higher risk of cognitive decline (n = 50). Pilot data analysis indicated a slight positive bias between model predictions and the clinical experts’ judgement (mean difference 0.98 units, limits of agreement from −0.83 to 2.78 units, reported to 3 s.f.). This analysis also revealed that the traffic light system, designed to indicate the risk of cognitive decline, can serve as a complementary source of information to enhance decision support. This is especially evaluated for its applicability in improving clinical decision-making for high-risk groups, where there is often insufficient data available regarding an individual’s cognitive status.

**CONCLUSIONS:** The study offers new insights into sleep and dementia, highlighting how age and sex differences manifest differently in typical ageing compared to dementia. We demonstrate the utility of leveraging sleep monitoring data and predictive analysis in identifying individuals who may benefit from further clinical evaluation and early disease-modifying interventions.

## 1 Introduction

There are approximately 57 million people living with dementia (PLWD) worldwide.^1^ It is estimated that three-quarters of individuals with dementia have not received a formal diagnosis, which significantly limits their access to treatment, care, and support. In low- and middle-income countries, this “treatment gap” is even more pronounced, reaching as high as 90%.^2^ As a consequence, people living with undiagnosed dementia are more likely to require emergency care and hospitalisations than their age-matched healthy counterparts.^3–5^ Despite this, dementia remains underdiagnosed across the globe, particularly in the earlier stages of its progression, where symptoms may instead be considered congruent with cognitive ageing.^2^ Even in the absence of disease-modifying treatment, early risk stratification for dementia has positive implications for managing comorbidities, mitigating safety risks, preventing complications, and supporting patients and their caregivers in planning for the future. Current screening methods, such as Positron Emission Tomography (PET), Magnetic Resonance Imaging (MRI) scans and fluid biomarker tests, are expensive, invasive, and unsuitable for large-scale screening. As such, there is an unmet need for more affordable, safe, and non-invasive tools that can reliably screen for dementia in the general population.

Age is a significant risk factor for dementia, with incidence increasing exponentially in older populations.^6^ Age is also associated with significant changes in sleep patterns.^7–9^ Older adults often experience shorter^10^ and more fragmented sleep, with increased nocturnal sleep disturbances.^11^ Moreover, older adults are more likely to experience multimorbidity (the co-existence of two or more chronic conditions^12^), with more than half of the world’s adult population over the age of 60 having multimorbidity.^13,14^ Multi-morbidity in older adults is also associated with self-reported sleep problems,^15–17^ which are often considered an outcome of multimorbidity but are also associated with a higher risk of experiencing symptoms of cardiopulmonary, vascular-metabolic and musculoskeletal conditions.^18^ In particular, changes in sleep physiology are also associated with an increased risk of dementia.^19,20^ In turn, dementia-related brain pathology changes have been associated with altered sleep patterns, with sleep disturbance, such as disruptions in slow-wave sleep or nocturnal awakenings, frequently emerging in the early stages of the disease and continuing throughout its progression.^21–23^ However, the exact causal relationship between sleep and cognitive decline remains unclear.

Recent works have used both electroencephalography (EEG) and polysomnography (PSG)-derived sleep data to predict age.^24–29^ Considering the association between age and changes in sleep patterns, the predicted sleep age offers a practical and readily measurable proxy for brain health.^29^ Sleep Age Index (SAI) is measured as the difference between a person’s chronological and predicted age.^24–29^ Higher SAIs have been statistically associated with a reduced life expectancy,^26,29^ as well as an increased risk of a variety of health conditions, including diabetes,^25^ HIV,^28^ hypertension,^25^ and dementia.^27^ Despite this, the use of SAI as an indicator of cognitive health has not yet been demonstrated as a predictor of dementia risk.

While these works have leveraged either EEG or PSG-derived sleep data, no studies have yet exploited remote monitoring of sleep in real-world settings to estimate age. PSG, a comprehensive assessment of sleep physiology, includes EEG, electrocardiography (ECG), electrooculography (EOG), chin and leg electromyography (EMG), and assessments of breathing effort and airflow.^29^ While PSG is widely regarded as the gold standard for diagnosing sleep disorders, its high running costs, intensive administrative requirements, and limited accessibility significantly constrain its capacity to generate broad clinical impact.^29,30^ Sleep EEG offers a more cost-effective and simpler alternative to PSG, but shares many of the limitations of using PSG, one of the main ones being how the unfamiliar environment required for data collection can alter participants’ typical sleep patterns.^31^ The advent of remote monitoring technologies has extended our ability to study sleep. Unlike PSG and EEG, remote monitoring technologies such as wearable devices and under-the-mattress sleep sensors^30^ enable the collection of sleep data in individuals’ natural sleep environments. However, wearable technologies usually have a short battery life, requiring regular charging and frequent removal.^32^ In studies involving populations where memory or dexterity may be impaired, this can limit the reliability of longitudinal monitoring. For this reason, we sought to assess the feasibility of using under-the-mattress sensors to study changes in sleep patterns associated with age and dementia.

This study presents a two-stage machine learning approach to identify dementia risk from SAI. The SAI scores are predicted using in-home nocturnal activity and physiology data collected by under-the-mattress sleep sensors. We have also developed a risk stratification method to improve the clinical applicability of our framework. The models have been trained and validated using two datasets: (1) a dataset comprised of 1,567 individuals from the general population, used here as a control, and (2) a dataset comprised of 105 individuals participating in an ongoing clinical study observing PLWD in their own homes. We also assessed the utility of our work as a tool to identify dementia risk by applying our approach to a high-risk population of 50 individuals in a pilot study. Figure 1 provides an overview of the three pre-processed datasets used in the training and evaluation of this approach. Our work (1) offers insights into how sleep changes with age, multimorbidity, and dementia, providing a foundation to guide discovery research (2) offers an early risk identification method and clinical decision support for earlier risk identification of dementia.

**Figure 1:**
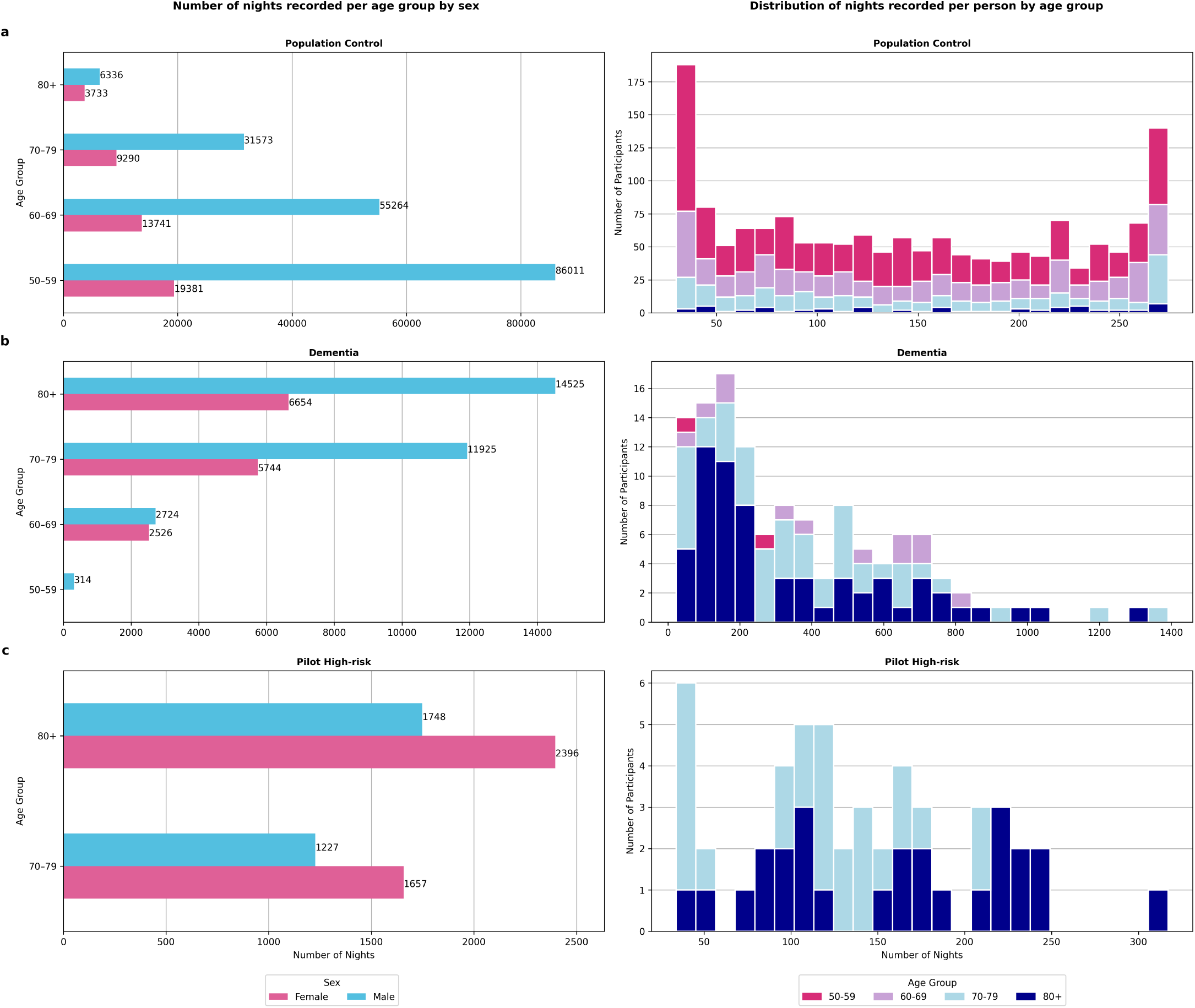
The composition of the pre-processed datasets from which the final processed samples were derived. The total number of nights recorded across age and sex (left-hand column), and the distribution of nights recorded by participant (right-hand column), where each bar represents the number of participants for which a given number of nights were recorded, illustrating the variability in counts across participants, and counts are grouped into 25 bins. Participants were grouped into four age bands. Coloured bars represent the four different age groups: 50-59, 60-69, 70-79, and 80+. a) Population Control cohort. b) Dementia cohort. c) Pilot High-risk cohort.

## 2 Results

This work included nocturnal activity and physiology data from three cohorts collected using under-the-mattress sleep sensors. After data processing, the Population Control cohort was comprised of 15,229 samples, collected from 1,567 participants (326 female, 1,241 male), the Dementia cohort was comprised of 3,140 samples collected from 105 participants (40 female, 65 male), and the Pilot High-risk cohort was comprised of samples 470 collected from 50 participants and their study partners (40 participants, 10 study partners; and 29 female, 21 male).

### Age estimation

We trained a regression model to estimate age using a training set comprised of 12,138 samples, collected from 1,253 participants (257 female, 996 male) derived from the Population Control cohort. We evaluated and compared the effectiveness of our model using three different regression models: Elastic Net, Huber, and Extreme Gradient Boosting (XGBoost). Table S9 in Supplementary Information Section S6 shows the results of the 10-fold cross-validation on all models tested. We compared model performance with and without sample weighting. We found the best-performing regression model was XGBoost with *L*1 regularisation, without sample weighting. While sample weighting tended to marginally improve performance on under-represented ages (as measured by the balanced mean absolute error, or BMAE) across all models, the overall regression performance (as measured by mean absolute error, or MAE, and mean squared error, or MSE) was almost always slightly worse compared to the unweighted model. Table 1 presents the model’s performance on both the validation and test data (definitions of evaluation metrics can be found in Supplementary Information Section S3).

**Table 1:** Mean (95% CI) % of MAE, MSE, and BMAE of the age estimation model on the different data splits. *MAE* Mean Absolute Error, *MSE* Mean Squared Error, *BMAE* Balanced Mean Absolute Error.

|  | MAE | MSE | BMAE |
| --- | --- | --- | --- |
| **Validation** | 5.97 (5.73 - 6.19) | 56.0 (52.0 - 59.7) | 6.59 (6.29 - 6.87) |
| **Test** | 5.52 (5.37 - 5.67) | 48.9 (46.2 - 51.6) | 7.37 (7.06 - 7.69) |

### How sleep metrics contribute to age prediction

SHapley Additive exPlanations (SHAP) were used to explain the individual predictions of the model. In regression tasks, SHAP plots show how much each feature contributes to the prediction for a given instance above or below the baseline (average predicted value, in this case, age). The magnitude of a SHAP value indicates how strongly a feature impacts a specific prediction, while the sign (positive or negative) indicates whether the feature increases or decreases the predicted age compared to the baseline, respectively. This demonstrates the key sleep metrics contributing to age prediction. Figure 2 shows a summary plot of SHAP values for the held-out test set from the Population Control cohort. The features are ordered with respect to their global importance. Figure S4 in Supplementary Information Section S7 shows a summary of the key sleep metrics contributing to age prediction in the Dementia cohort.

**Figure 2:**
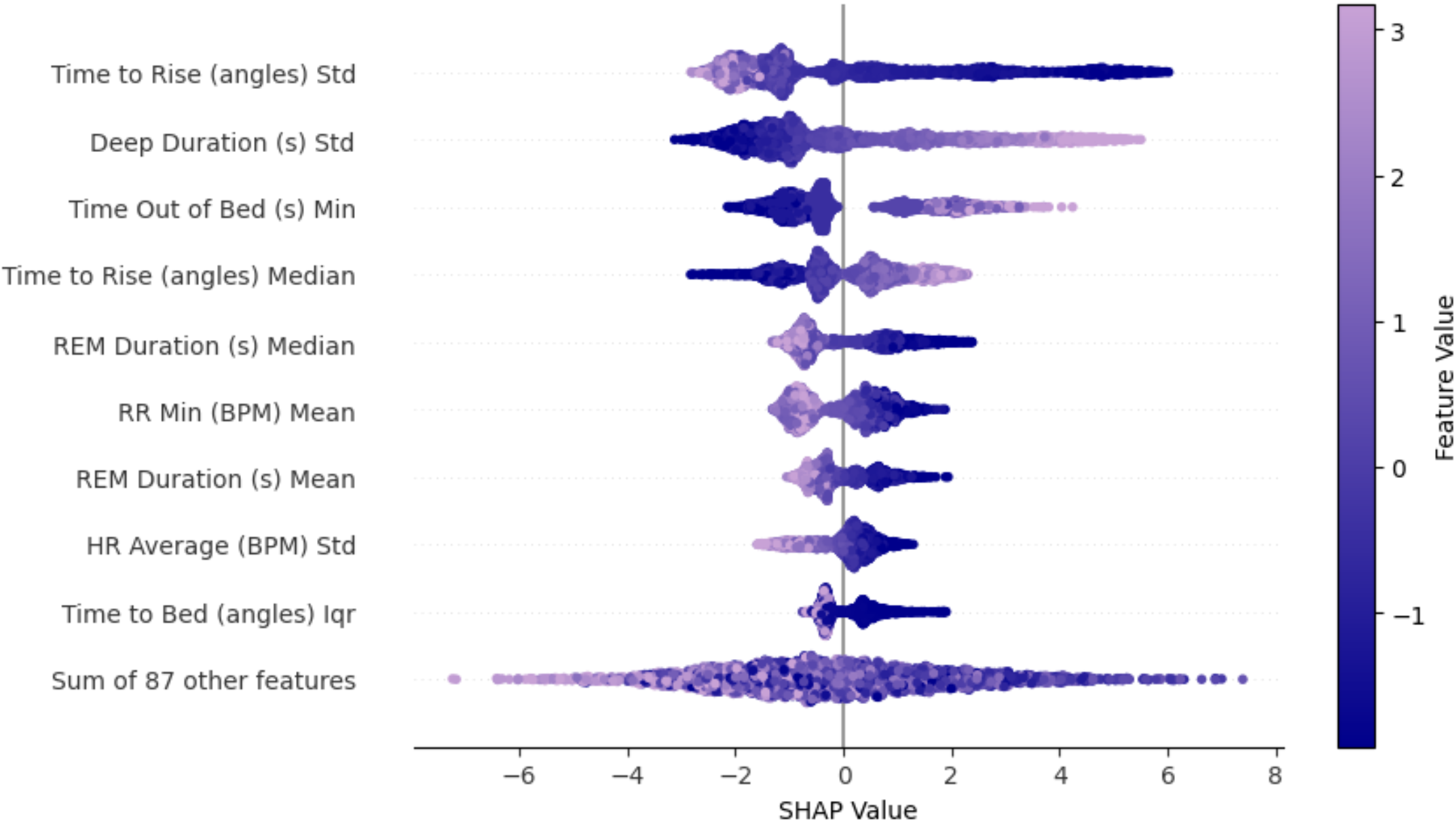
SHAP values. The feature importance for the top 10 most important features, as calculated on the held-out test set using SHAP. The corresponding feature values are coloured according to the value of that feature (high values shown in lilac and low values in dark blue), whilst the position along the x-axis represents the contribution that particular value made to the prediction.

### Calculating SAI

To calculate the SAI, we applied a three-step procedure (see Figure S1 in Supplementary Section S1 for a visualisation). In Step 1, we obtain predicted sleep ages for each cohort (Population Control test set, Dementia, and Pilot High-risk) using our pre-trained age estimation model. The pre-trained model used the control dataset for training and validation. We compared the performance of our age estimation model across female and male participants, within both the Population Control and Dementia cohorts (see Table S10 in Supplementary Information Section S6). In Step 2, we adjust these predictions based on age-group-weighted mean differences between the sleep and chronological age in the Population Control test set (n = 314 participants, 69 female, 245 male). To train the age estimation model, we used broader age bands (50–59, 60–69, 70–79, and 80+) for stratification and sample weighting to maintain training stability and reduce data fragmentation. Narrower age bands (50–54, 55–59, 60–64, 65–69, 70–74, 75–79, and 80+) were employed in step 2 to calculate age-group-weighted means that aligned better with clinically observed age-related changes in sleep patterns. These means were then subtracted from the predicted sleep ages (computed in step 1) across all three datasets, based on the participant’s age group. In step 3, we computed the SAI for each participant as the difference between the age-group-adjusted sleep age and chronological age.

A statistically significant difference in SAI means was observed between the Dementia cohort (n = 3,140, mean = –4.21, SD = 5.86) and the Population Control cohort (n = 3,091, mean = 1.34, SD = 4.55) (t = –41.7, p ¡ 0.001), all reported to 3.s.f., supporting the use of SAI as a predictive feature in our dementia risk screening model. Figure S5 in Supplementary Information Section S7 visualises the results of a quantitative analysis comparing SAI with severity of cognitive impairment for individuals in the Dementia cohort. Severity classifications follow the guidelines of the National Institute for Health and Care Excellence^33^ (see Table S3 in Supplementary Information Section S2.

### Dementia risk prediction

We trained and evaluated a binary classification task to predict dementia risk by differentiating between the SAI of an individual from the Dementia cohort from that of an individual from the Population Control cohort. In total, our dataset consisted of 6,231 samples, collected from 419 participants (109 female, 310 male). Our training set was comprised of 4,927 samples, collected from 334 participants (79 female, 255 male). We evaluated and compared the effectiveness of two different classification models: Logistic Regression and XGBoost. Table S11 in Supplementary Information Section S8 shows the results of the 10-fold cross-validation on all models tested. We also compared model performance with and without sample weighting. We found the best-performing classification model was Logistic Regression with *L*2 regularisation, without sample weighting. Table 2 presents the model’s performance on both the validation and held-out test data (definitions of evaluation metrics can be found in Supplementary Information Section S3).

**Table 2:** Mean (95% CI) % of sensitivity, specificity, and area under the precision-recall curve of the dementia risk prediction model on the different data splits. These results are reported before and after stratification is performed.

Before Stratification
|  | Sensitivity | Specificity | AUC Precision-Recall |
| --- | --- | --- | --- |
| **Validation** | 68.8 (61.2 - 75.9) | 71.5 (69.0 - 73.8) | 78.8 (71.7 - 85.3) |
| **Test** | 69.2 (65.5 - 72.7) | 62.8 (59.1 - 66.5) | 76.8 (73.7 - 79.7) |

After Stratification
|  | Sensitivity | Specificity | Precision |
| --- | --- | --- | --- |
| **Validation** | 79.9 (68.7 - 91.0) | 86.7 (76.1 - 93.7) | 86.7 (74.7 - 94.9) |
| **Test** | 75.7 (71.4 - 79.9) | 74.7 (69.2 - 80.0) | 82.1 (78.0 - 86.1) |

### Risk stratification

We developed a traffic light system to stratify the dementia risk based on the model’s confidence in its predictions. We varied stratification thresholds to balance sensitivity and specificity with the number of samples stratified into the Red and Green groups. In this work, we select thresholds [0%, 30%], [30%, 70%], and [70%, 100%] (following interval notation) for the ‘Green’, ‘Amber’, and ‘Red’ groups, respectively. Table 2 shows the model performance after stratification when evaluating the predictions on the ‘Green’ and ‘Red’ groups. Figure 3 reports the distribution of the model’s predictions by group (including the ‘Amber’ group) on the held-out test set (n = 1,304, p = 85). We provide an analysis of model reliability and calibration in Supplementary Information Figure S6 in Supplementary Information Section S9. We also compare model performance between sex and across age groups (see Supplementary Information Tables S12 and S13, respectively, in Supplementary Information Section S11).

**Figure 3:**
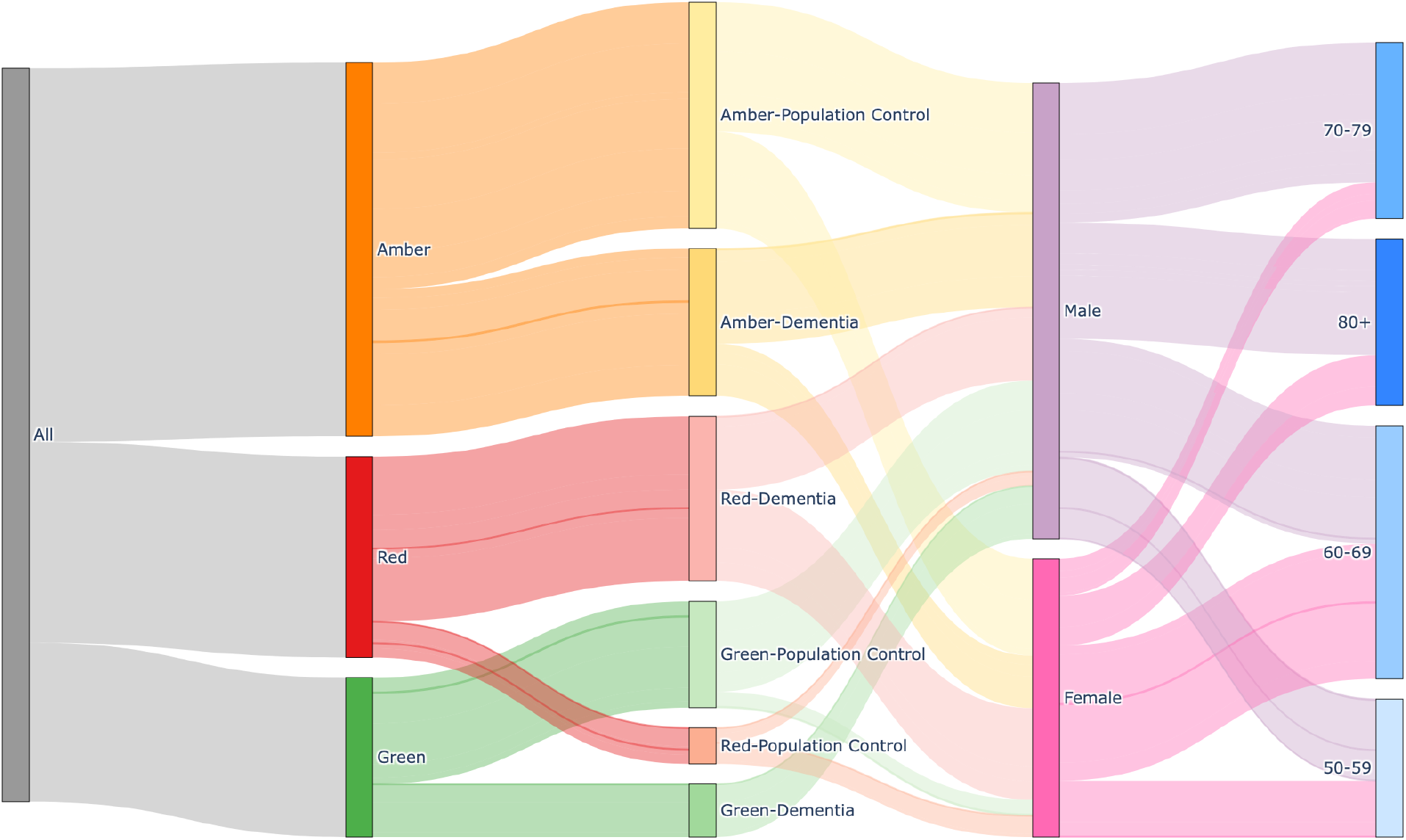
Sankey diagram showing stratification breakdown of risk prediction model. Analysis of stratified groupings produced by evaluating the final classification model on the held-out test set using optimised thresholds. ‘Green’ represents the samples that the model confidently predicts are from an individual in the Population Control cohort, ‘Amber’ represents the samples that the model is not confidently predicting, and ‘Red’ represents the samples that the model confidently predicts are from an individual in the Dementia cohort. Each group is further broken down by the true labels, then by the sex of the participant from which the sample came, and finally by age (using the four age bands for the sake of interpretability). View an interactive version of the figure here: *https://github.com/tmi-lab/Sleep-Age-Dementia/blob/main/sankeydiagram.html*.

### Pilot study in a high-risk population

To assess the predictive accuracy of our framework, we ran a pilot study within the NHS. We applied our classification model (and stratification thresholds) to the SAI for the Pilot High-risk cohort, a real-world high-risk population of individuals who may plausibly have undiagnosed dementia or go on to develop the condition.

Following dementia risk prediction on the held-out test set (n=1,304, p=85), we calculated the age-group weighted mean dementia risk score using the narrower age bands (50 - 54, 55 - 59, 60 - 64, 65 - 69, 70 - 74, 75 - 79, 80+) for all known positive cases of dementia. We then subtracted these values from the dementia risk scores predicted for each sample in the Pilot High-risk cohort dataset (n=470, p=50). This allowed us to estimate similarity to diseased individuals by looking at how closely the dementia risk score of an individual in the Pilot High-risk cohort looks compared to the dementia risk scores of their age-matched counterparts in the Dementia cohort. By controlling for age, we are also able to correct for demographic data drift.

Using the age-group-adjusted difference between dementia risk scores, alongside stratification, we reassigned participant samples into three groups: ‘Red’, ‘Amber’, and ‘Green’, which represent ‘Highly Likely’, ‘Somewhat Likely’, and ‘Not Likely’ risks of cognitive decline, respectively. Samples were reassigned using the following rules:

- If group == `‘Red’` and `diff` ≥ 0, return `‘Highly Likely’`
- If group == `‘Red’` and `diff` < 0 or group == `‘Amber’` and `diff` ≥ 0, return `‘Somewhat Likely’`
- Otherwise, where group == `‘Amber’` and `diff` < 0 or group == `‘Green’`, return `‘Not Likely’`

On applying these criteria for reassignment to our Pilot High-risk cohort, we were able to generate profiles for each participant based on the model’s confidence that they may be experiencing cognitive decline over time. To evaluate the utility of our pipeline as a tool for identifying dementia risk, a subset of this cohort (the study participants, n = 40) underwent clinical assessment. Two clinicians (experts in dementia) received a list of the participant IDs included in our analysis, but were blinded to the model’s output to reduce potential bias. We measured the agreement between the predictions for each participant and the ground truth determined by clinical investigation. For the clinical investigation, the experts used the Addenbrooke’s Cognitive Examination-III (ACE-III) scores and electronic health records of each participant in the pilot study. Figure 4(a) shows how dementia risk as predicted by our model varies over time in individuals from the high-risk population. We have coloured the bars by the stratification groups to which participants were reassigned based on their adjusted risk scores. To further protect individual privacy, we replaced the original anonymous participant IDs with a new set of randomly generated numbers. We have further annotated the plot with the clinical labels given by each clinician (an expert in dementia) to each participant during re-assessment. Figure 4(b) is a Bland-Altman^34^ plot visualising the level of agreement between the model’s predictions and expert annotations. The pilot data analysis indicated a slight positive bias between the model’s predictions and the clinical ground truth (mean difference 0.98 units, with limits of agreement from −0.83 to 2.78 units (mean difference ± 1.96 SD), all reported to 3.s.f.).

**Figure 4:**
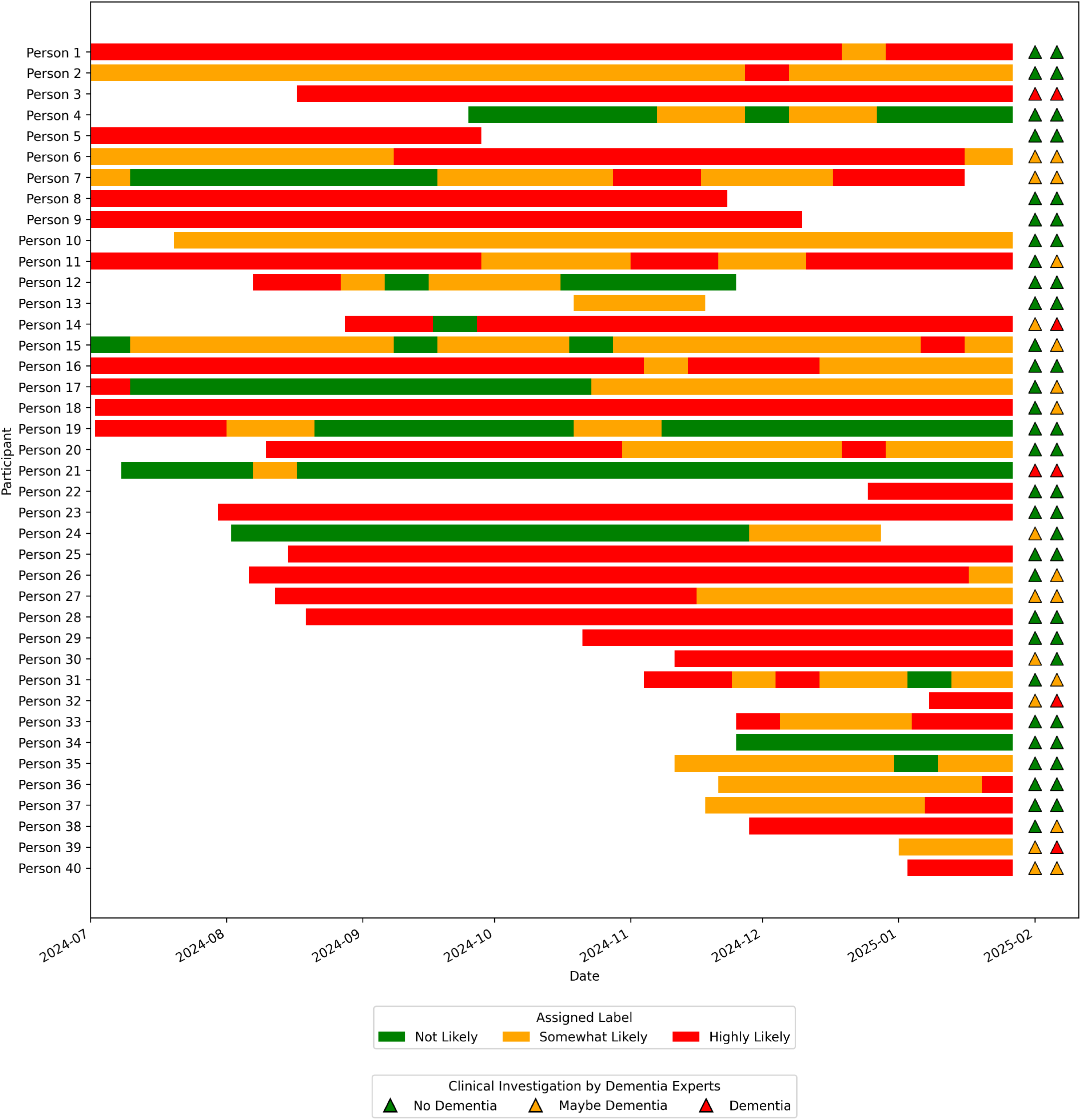
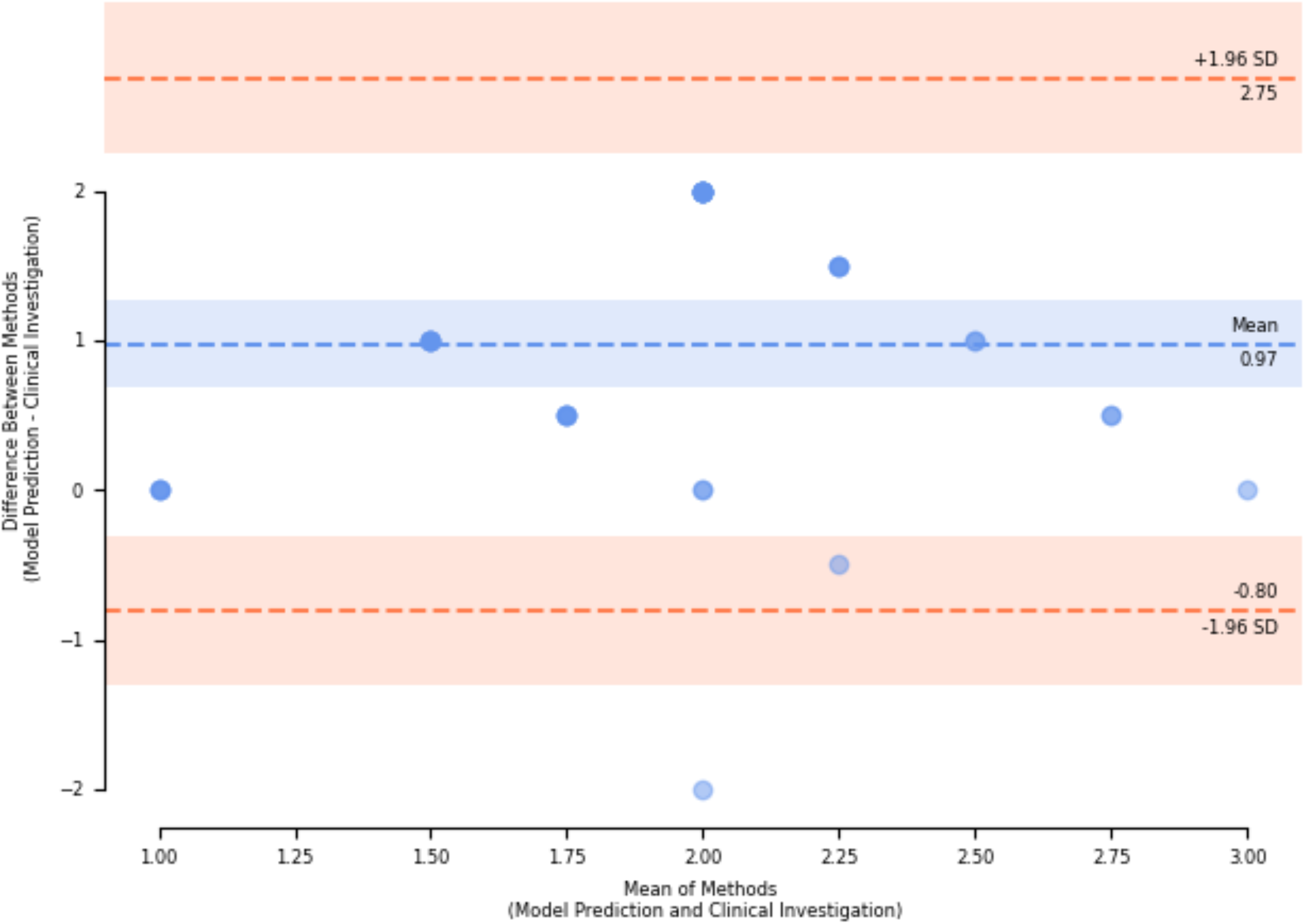
**(a)** Dementia risk over time for individuals in the Pilot High-risk cohort. Bar colours correspond to the predicted likelihood that an individual might have dementia (the assigned label): ‘Not Likely’ is shown in green, ‘Somewhat Likely’ in amber, and ‘Highly Likely’ in red. Triangle colours correspond to the clinical label given to each participant during reassessment: ‘No Dementia’ is shown in green, ‘Maybe Dementia’ in amber, and ‘Dementia’ in red. Each triangle represents the clinical ground truth determined by a single expert. **(b)** Bland-Altman plot showing agreement between the model’s predictions and expert annotations. Each point represents a paired measurement. From the 40 participants, we derive 11 unique paired measurements. The mean difference (bias) is represented by a dashed blue line, while the lower and upper limits of agreement (mean difference ± 1.96 SD) are represented by dashed red lines.

Finally, after accounting for within-participant variability using linear mixed-effects modelling, no statistically significant association was found between the adjusted risk scores and ACE-III scores (*β* = 0.05, sd = 0.03, z = 1.49, p = 0.14), reported to 3.s.f. Moreover, no statistically significant association was found between either the adjusted risk scores and PHQ-9 scores (*β* = −0.05, sd = 0.06, z = −0.94, p = 0.35) or the GAD-7 scores (*β* = −0.03, sd = 0.05, z = −0.60, p = 0.55), respectively, reported to 3.s.f.

## 3 Discussion

Sleep patterns undergo significant alterations with age. These alterations are reflective of changes in the broader neurological and physiological processes that occur during ageing. In recent years, the concept of leveraging sleep age, a biomarker derived by estimating chronological age using sleep features, has emerged as a promising index by which to measure brain health.^24–29^ An increase in predicted sleep age compared to chronological age has been associated with increased mortality and morbidity, including neurodegenerative conditions and cognitive decline, suggesting its potential in monitoring and early disease detec-tion.^25–29^ The primary aim of this study was to predict dementia risk using SAI derived from remote sleep monitoring data. Our framework is designed to be used as a risk screening tool to assist clinical decision-making. Our approach uses explainable machine learning models with traceable decision-making processes, ensuring our model remains a practical aid for clinicians.

### Two-stage approach

We present a two-stage machine learning approach for estimating dementia risk using SAI derived from remote sleep monitoring data. We considered several regression models for age estimation and found that XGBoost attained the top performance (MAE of 5.52 (95% CI: 5.37 - 5.67), MSE of 48.9 (95% CI: 46.2 - 51.6), and BMAE of 7.37 (95% CI: 7.06 - 7.69)). This performance was comparable to other age estimation models trained on EEG and PSG data,^24,25^ including those using deep neural networks.^29^

We next considered two common classifiers for age estimation and found that Logistic Regression attained the top performance (sensitivity of 69.2% (95% CI: 65.5% - 72.7%) and specificity of 62.8% (95% CI: 59.1% - 66.5%). The ratio of positives (samples from individuals in the Dementia cohort) to negatives (samples from individuals in the Population Control cohort) in the test set is 1 : 1.01. We then transformed dementia risk scores into three tuneable groups, allowing us flexibility in the management of model accuracy and uncertainty. Post-stratification, the performance on the ‘Green’ and ‘Red’ groups was improved, achieving a sensitivity of 75.7% (95% CI: 71.4% - 79.9%), and a specificity of 74.7% (95% CI: 69.2% - 80.0%) (see Supplementary Information Section S2 for definitions of evaluation metrics). Grouping the classification model’s predictions based on the dementia risk score also increased accuracy in each sex and across the four age bands, as shown in Tables S12 and S13 in Supplementary Information Section S11. A breakdown of these predictions can be seen in Figure 3.

### Pilot study

We conducted a Pilot study applying our model to a high-risk population of multi-morbid individuals. Figure 4(b) visualises an analysis of the agreement between the model’s predictions and the ground truths established by dementia experts. This analysis indicated a slightly positive bias (0.98) overall and moderate agreement between the model and expert annotations. Specifically, there was good agreement in clear cases of dementia or no dementia, but slightly poorer agreement for more ambiguous cases. These findings suggest that our model is reliable for distinguishing between PLWD and those without, but somewhat less reliable for borderline cases. We also present an evaluation of the relationship between the adjusted risk scores and the ACE-III, the Patient Health Questionnaire-9 (PHQ-9) and Generalised Anxiety Disorder Questionnaire-7 (GAD-7) scores, respectively, in the Pilot High-risk cohort. Accounting for within-subject variability, linear mixed-effects modelling found no statistically significant associations across all three pairings. Self-reporting can introduce subjectivity and potential inaccuracies in responses.^35^ This bias can compromise data reliability and this may contribute to the lack of statistically significant associations observed in this study. Overall, these findings serve as preliminary evidence of the feasibility for use of our framework as a risk screening tool in larger clinical trials. Subsequent analysis also reveals that the traffic light system, designed to indicate the risk of cognitive decline, can serve as a complementary source of information to enhance decision support, which is especially useful for high-risk groups, where there is often insufficient information regarding an individual’s cognitive status.

### Explainability

Our approach uses SHAP analysis to reveal how individual sleep metrics contributed to age prediction. Based on analysis of held-out test data from the Population Control cohort, we observe that higher variability of time to bed and time to rise were associated with lower predicted ages. This aligns with previous findings showing that younger adults’ sleep-wake schedules tend to be more irregular, impacted by changing social and working schedules that make it harder to maintain a consistent sleep routine compared to older adults (*>* 65 years of age).^36^ Likewise, higher variation in heart rate and higher minimum respiratory rate are also associated with lower age predictions. The latter suggests that low respiratory rates contribute to higher age predictions. Our analysis also highlighted that high variability in deep sleep duration and less REM sleep were associated with higher predicted ages. In particular, disruptions in slow-wave (deep) sleep have been linked to the pathophysiology seen in neurodegenerative disease and have been suggested as a potential biomarker for early diagnosis.^37–39^ Finally, more time spent out of bed between going to bed and rising was also associated with higher predicted age, suggesting older adults were experiencing more frequent awakenings. This finding is in agreement with the literature.^40^

In contrast to previous reports,^27^ we found PLWD tended to be associated with negative SAI (they were predicted younger) after adjusting for age, whereas positive SAI scores were more commonly observed in the Population Control cohort (they were predicted older). Sleep and ageing are tightly coupled in healthy adults; however, this relationship may be disrupted in neurodegeneration.^7^ While dementia is associated with sleep fragmentation and reduced REM and slow-wave sleep,^21–23^ these represent pathological changes that do not necessarily align with typical age progression. Our model was trained using a cohort from the general population. As such, it has learned sleep patterns associated with typical ageing, explaining our model’s misinterpretation of these abnormal dementia-related patterns with respect to chronological age. For example, the fragmentation of sleep in dementia may cause increased restlessness, mimicking the more irregular sleep patterns of younger adults,^36^ justifying why a PLWD might experience higher time to bed or time to rise variability. Likewise, where disruptions in the slow-wave sleep of older adults may lead to increased night-to-night variability, a PLWD might experience consistently low slow-wave sleep, reducing the variability observed in these individuals.

### The effect of the chosen technology

We leveraged under-the-mattress sleep sensors to collect personalised longitudinal data from people in the comfort of their own homes. The sensor used in this study has been validated to detect total sleep time, sleep efficiency and wake after sleep onset in a range of studies,^41–43^ including for people with sleep disorders such as sleep apnoea,^44^ and is reported to have comparable performance to wearable sleep trackers.^45^ In addition to providing a scalable, low-cost approach for data collection in naturalistic settings (unlike in PSG or EEG studies), under-the-mattress sleep sensors have the potential to reduce data loss compared with wearable devices.^45^ In studies focusing on age-related diseases where individuals may be experiencing cognitive or functional impairment, under-the-mattress sleep sensors boast a particularly important advantage.

### Reliability and fairness

In Supplementary Information Section S9, we discuss the model reliability and calibration and find that, while our model is reasonably well calibrated, it still experiences some non-linear miscalibration when predicting dementia. This motivates the application of our stratification thresholds to the model’s dementia risk scores, as it allows us to balance model accuracy with the number of ‘Red’ flags; however, in deployment, the trade-off between sensitivity and specificity should be re-evaluated to ensure the groups remain well-calibrated for the cohort being studied. Considering sex (see Supplementary Information Table S10), we see that while our age estimation model had similar performance for both sexes, female participants were more likely to be predicted as having dementia (see Supplementary Information Table S12). Considering age (see Supplementary Information Table S13 in Supplementary Information Section S11 for a preliminary analysis of model performance across each age group), we see that our model performs well between the ages of 50-59 and on participants over 80, reasonably well between the ages of 60-69, but has a tendency to yield false negatives between the ages of 70-79, while we also note a large imbalance in the number of samples from each cohort within the age ranges of 50-59 and 80+, respectively. When deploying this as a risk screening tool, the proportion of labels might impact these metrics. For this proof-of-concept study, we have trained a single risk prediction model to prevent overfitting in the model due to insufficient data; however, another direction of this work could look to train separate models to make predictions on each of the four age bands, as well as for male and female participants.

In addition, in Supplementary Information Section S12, we present an evaluation of the effect of seasonality on both our age estimation and our risk prediction. While there was no meaningful effect on the final classification decisions, in both the Population Control and Dementia cohorts, differences in SAI were observed between the Winter and Summer months. Further work would benefit from examining the potential impact of environmental factors such as temperature and daylight hours on sleep metrics. Moreover, validating this framework in regions with different seasonal patterns might further improve generalisation and robustness.

### Limitations of the data included

Our prediction model did not include information on comorbidities or medications, factors which may influence the risk of dementia. Multimorbidity is also associated with a higher prevalence of sleep problems, increased nocturnal disturbances and altered sleep-wake routines.^15–17^ The motivation for this work was to develop a framework for identifying high-risk, clinically complex populations for dementia. A future direction of this work is to incorporate these variables to tailor the model, thereby more accurately differentiating between individuals with and without dementia, as well as in the context of other conditions and treatments, enhancing predictive performance and clinical relevance.

### Conclusions

Here, we provide preliminary evidence for a scalable framework to identify the risk of dementia in the general population, which, with further testing, has the potential to transform the field of dementia care. To the best of our knowledge, while previous studies have used EEG sleep data to predict dementia directly,^46^ this is the first study that uses remote sleep monitoring data to predict dementia, using SAI as a proxy measure for cognitive health. The proposed pipeline takes advantage of the inherent interpretability of machine learning models to develop a transparent and explainable framework, designed to enhance clinical decision-making. In addition, utilising stratification techniques enables us to calibrate our classification model for improved performance in clinical settings. Furthermore, SHAP analysis (such as in Figure 2 presents explainable results that allow researchers and clinicians to explore why the algorithm has made a particular prediction and better understand the difference between how ‘normal’ ageing and dementia affect sleep. By identifying sleep patterns associated with cognitive decline from those associated with typical ageing, our framework can be used to guide discovery research. Finally, we conducted a prospective pilot study on a high-risk population of multi-morbid individuals to determine the model’s usefulness as a tool for predicting dementia risk.

This approach can be scaled rapidly. The use of this framework has the potential to markedly improve the quality of life for PLWD, supporting earlier diagnosis and the development of more timely care plans.

## 4 Methods

### Data use and study design

We include sleep data collected from participants across three study cohorts (each with a different set of study objectives and population cohort) using an under-the-mattress sleep sensor: a dataset obtained from users of the Withings sleep mat monitoring device within the general population, referred to in this paper as the Population Control cohort; a dataset obtained from the Minder Integrated Health Management Study,^47^ the UK Dementia Research Institute (UK DRI) Care Research & Technology (CRT)^48^ Centre’s ongoing longitudinal study of PLWD in the United Kingdom, which began in 2018, referred to in this paper as the Dementia cohort; and a dataset obtained via the Resilient^49,50^ study, the Surrey and Borders Partnership NHS Foundation Trust’s ongoing feasibility study collecting in-home monitoring data from a cohort of older adults (65+) diagnosed with multi-morbidity at the risk of cognitive decline and their study partners (who provide essential care and support at home of the main study participants), referred to in this paper as the Pilot High-risk cohort.

#### Population Control

The population control cohort comprises individuals from the general population using the Withings sleep mat monitoring device.^51^ This dataset includes over 1 million nights of sleep from June 2020 to March 2021 across 5,574 participants in Europe.

#### Dementia

Eligible study participants for the Minder Health Management Study include adults older than 50 years of age who are clinically diagnosed with dementia, MCI, frailty, stroke, or traumatic brain injury. Individuals with severe psychiatric symptoms (including the presence of active suicidal thoughts or severe mental health conditions such as depression, anxiety, psychosis, and agitation), severe sensory impairment, or those receiving treatment for terminal illness are excluded from the study. This dataset includes over 110,000 nights of person-data across 160 participants collected from the 1st of December 2018 to the 3rd of April 2025.

#### Pilot High-risk

Eligible study participants include individuals aged 65 or older with a clinical diagnosis of at least two chronic conditions associated with an increased risk of dementia. These included, but were not limited to, arthritis, chronic kidney disease, heart disease or heart failure, depression and other mental health disorders, diabetes, hypertension, liver disease, and stroke. Participation was open to individuals with or without a study partner. Exclusion criteria include severe mental health conditions (such as psychosis, severe depression, agitation, or anxiety), profound sensory impairments, current treatment for a terminal illness with a life expectancy of less than six months or in the final year of life, and the inability to provide informed consent. The final eligibility decisions were made by the principal investigator on a case-by-case basis. This dataset includes over 14,000 nights of person-data collected between the 25th of August 2023 to the 25th of February 2025 across 74 participants and their study partners (55 participants, 19 study partners).

### Ethical approval

The Population Control dataset was provided by Withings (https://www.withings.com) under a data-sharing agreement for research with Imperial College London. Withings is certified in ISO 27001 and Health Data Hosting (HDS) which comply with the same level of security as health professionals. Users of the Withings under-the-mattress sleep sensor consented to their data being used as part of the Withings research programme^52^ and may withdraw their consent at any time. The data used in this work was fully anonymised before sharing, so re-consent was not required. Withings data collection is in compliance with the GDPR, with their data privacy policy ((https://www.withings.com/uk/en/legal/privacy-policy)) citing “legitimate interest” as one of the lawful biases for processing personal data, in this case, Withings processes non-identifying data, to improve research. Information governance and impact assessment approval for storing and processing data shared by Withings was obtained from the Surrey and Borders Partnership NHS Foundation Trust (reference number: 20201009-DRI).

The Minder Health Management Study is registered with the National Institute for Health and Care Research (NIHR) in the United Kingdom under the Integrated Research Application System (IRAS) registration number 257561 and received ethical approval from the London-Surrey Borders Research Ethics Committee; TIHM 1.5 REC: 19/LO/0102. Upon joining the study, all participants and their study partners provided written informed consent to participate in the study and for their data to contribute to publications. If deemed unable to do so, consent was obtained from either a participant’s legal representative(s) or power of attorney holder(s) after discussions with family members and clinicians to ensure that participation in this study was in the participant’s best interest. Approval to collect data from the under-the-mattress sleep sensor in the Minder Health Management Study was granted by a clinical IRB panel.

The Resilient study received ethical approval from the London-Surrey Borders Research Ethics Committee and the Health Research Authority, and was formally registered on the Integrated Research Application System (IRAS) under reference number 321104. All participants provided written informed consent prior to enrolment. Participants received a participant information sheet that explained how their personal data would be collected, used and stored in compliance with the General Data Protection Regulation (GDPR). Individuals who were deemed to lack capacity at baseline were excluded from the study.

For both the Minder Health Management Study and the Resilient study, capacity to consent was assessed in accordance with Good Clinical Practice (GCP),^53^ as outlined in the UK Policy Framework for Health and Social Care^54^ Research and the Mental Capacity Act 2005.^55^

### Data pre-processing procedures and characteristics

To control for age and ensure a more meaningful comparison between datasets, we select only those individuals aged 50 years and older from the Population Control dataset. All datasets underwent identical pre-processing. Aggregated by participant, data was cleaned, and missing dates were imputed using a rolling mean with a 7-day window in which a minimum of 5 days needed to be non-zero values. All remaining dates with missing data were removed. To maximise the information in the data, we generated two additional features (‘Time to Bed (angles)’ and ‘Time to Rise (angles)’). The dataset was comprised of a total of 16 features (‘Time to Bed (angles)’, ‘Time to Rise (angles)’, ‘Time in Bed (s)’, ‘Time Out of Bed (s)’, ‘Bed Time Period (s)’ (total time in bed), ‘Rapid eye movement (REM) Duration (s)’, ‘Light Duration (s)’, ‘Deep Duration (s)’, ‘Awake Duration (s)’, ‘Time Snoring (s)’, ‘Heart rate (HR) Average (beats per minute or BPM)’, ‘HR Minimum (BPM)’, ‘HR Maximum (BPM)’, ‘Respiratory rate (RR) Average (breaths per minute or BPM)’, ‘RR Minimum (BPM)’, ‘RR Maximum (BPM)’). For the purposes of training our age estimation model, we grouped participants into one of four age bands (50-59, 60-69, 70-79, 80+) based on their chronological age. We then filtered for participants who did not have at least n = 30 days of consecutive-day data. We then derived n = 30-day arrays of data for each participant using a rolling window of 10 days. This approach allowed us to maximise data usage, minimise information loss between data periods, and improve our ability to detect more gradual changes. Our choice to select n = 30 days based on previous work by this group^56^ was further validated during preliminary evaluations of our age estimation models (see Supplementary Information Section S6). We calculate statistics (the mean, median, minimum, maximum, standard deviation and interquartile range of each feature) for each array of participant data. Sex is included as a confounding variable, with the data transformed into a numeric format using one-hot encoding. The resulting processed data was comprised of a total of 98 features.

Demographics of the three datasets are provided in Table S1 in Supplementary Information Section S2, reporting both the original dataset demographics and those of the processed versions used in this study. Male and female assignments were self-declared by participants. We also include information on the primary diagnoses and Mini-Mental State Examination (MMSE) scores used to assess the severity of cognitive impairment for the Minder Health Management Study participants on entry into the clinical study in Table S2 in Supplementary Information Section S2. Additionally, we include the top five most common diagnoses and medications for participants in the Resilient study (see Tables S4 and S5 in Supplementary Information Section S2, respectively), as well as ACE-III scores used to assess the severity of cognitive impairment on entry into the clinical study, along with the respective severity classifications^57,58^ (see Table S6 in Supplementary Information Section S2). Additionally, we report the PHQ-9 and GAD-7 scores, as well as the respective severity classifications of depression and anxiety^59,60^ for study participants at the time of entry into the clinical study (see Tables S7 and S8 in Supplementary Information Section S2).

### Analysis platform

All analyses were performed on a secure computing environment at Imperial College London in Python version 3.13.2 environment. The Pandas^61^ (version 2.2.3), NumPy^62^ (version 2.1.3), SciPy^63^ (version 1.15.2), statsmodels^64^ (version 0.14.4), and Scikit-Learn^65^ (version 1.6.1) packages formed much of our pipeline. All visualisations were plotted using Matplotlib^66^ (version 3.10.1), Seaborn^67^ version (0.13.2), pyCompare^68^ version (1.5.4), and SHAP (SHapley Additivie exPlanations), including the high-speed Tree Explainer algorithm^69,70^ (version 0.47.0).

### Statistical Analyses

Prior to deriving our data arrays for age estimation, we performed statistical analyses on individuals from the Population Control cohort (the processed dataset) with at least 30 consecutive days of data. We conducted between-subjects parametric (analysis of variance, or ANOVA) and non-parametric (Kruskall-Wallis) tests to investigate the relationship between age group and nocturnal activity and physiological data. We modelled the fixed effects of age. Post-hoc, Welch t-tests were used to identify which groups differed from each other. Holm-Bonferroni was used to correct for multiple comparisons. See Figure S2 in Supplementary Information Section S4 for a visualisation of these comparisons. These preliminary analyses verified the validity of using these features to predict sleep age. After estimating the sleep age, we first calculated the Cohen’s d statistic to measure model performance between sexes for both the Population Control and Dementia cohorts. We then conducted a between-subjects Welch’s t-test (parametric) to investigate the relationship between the cohort to which the individual belonged and the SAI. We also used a one-way between-subjects ANOVA to investigate the relationship between severity of cognitive impairment and SAI, followed up by post-hoc Tukey’s HSD (Honestly Significant Difference) tests. Finally, we used a one-way between-subjects ANOVA to investigate the effect of seasonality on both our age estimation and dementia risk prediction models, followed up by post-hoc Tukey’s HSD tests where necessary. Unless otherwise stated, we use standard significance reporting (p *<* 0.0001 denoted as ***, p *<* 0.001 denoted as **, p *<* 0.05 denoted as *, and p *<* 0.1 denoted as). For our pilot study, we conducted a Bland-Altman analysis^34^ to measure the strength of agreement between our model’s predictions and the ground truths established by dementia experts in the Pilot High-risk cohort. Then, using the statsmodels^64^ package (version 0.14.4), we also conducted linear mixed effects modelling to examine the relationship between the adjusted risk scores and measures of cognitive impairment (ACE-III), depression (PHQ-9) and anxiety (GAD-7) in the Pilot High-risk cohort as the respective outcome variables. We also accounted for within-subject variability by specifying random intercepts for each participant.

### Framework for age estimation

Our age estimation models were trained and evaluated on the processed Population Control dataset (n = 1,567 participants assumed to be representative of typical ageing). To ensure generalisability, this dataset was split into training and testing subsets in an approximate 80:20 ratio, split by ID, with stratification applied in the four age bands to preserve age distribution across the training and test sets. The training data represented 80% of the participant IDs (n = 1,253 participants) and the testing data represented the remaining 20% (n = 314 participants). During model development and whilst optimising model hyperparameters, validation sets were produced by splitting the training data (by participant ID) using stratified k-fold cross-validation. See Supplementary Information Figure S1 ‘Age estimation’ in Supplementary Information Section S1 for a visualisation of this evaluation. Stratification ensured that each fold contained a balanced representation of each age group compared to the whole training dataset, which is particularly important here due to the imbalance in the age group distributions. Finally, we used mean absolute error, mean squared error, and balanced mean absolute error to measure model performance (for definitions of metrics, see Supplementary Information Section S3).

#### Age estimation models

For predicting sleep age, we tested three commonly used machine learning models for time-series regression: Elastic Net,^71^ Huber,^72^ and XGBoost.^73^ These models represent a diverse and robust mixture of models by which to benchmark performance and get insights into feature importance. Hyperparameter optimisation was performed using k-fold cross-validation (k = 10), with the model producing the lowest mean absolute error on validation data selected for the final analysis. See Supplementary Information Figure S1 ‘Age estimation’ in Supplementary Information Section S1 for a visualisation of this evaluation. Supplementary Information Section S5 contains information on the decisions made regarding each step of the regression pipeline. Furthermore, given the imbalanced distribution of ages in the Population Control dataset, we used this evaluation to explore sample weighting as a strategy to mitigate potential bias towards overrepresented ages. We applied sample weighting using the discretised age groups, assigning weights inversely proportional to bin frequency.

We initially selected n = 30 days to pre-process our datasets, and we used this validation technique to evaluate model performance using statistics derived from 30, 60 and 90-day arrays of data, respectively. While statistics derived from 90-day arrays often provided slightly better performance across models, with and without sample weighting, (see Table S9 in Supplementary Information Section S6), the improvement was too marginal compared to the prediction delay. 30-day windows allow for more frequent and timely updates, key for clinical monitoring. Moreover, using a shorter time window increases the number of usable data points, improving the robustness and generalisability of the model, with the 30-day window resulting in nearly 185% more samples, helping to reduce overfitting (see Figure S3 in Supplementary Information Section S5). The selection of n = 30 days resulted in the final sample and participant numbers for the processed datasets as presented in this paper.

### Framework for predictive analysis

Our risk prediction model was trained and evaluated on a dataset of the learned SAI from both the held-out test set of the Population Control dataset and the Dementia cohort. To allow for research interpretability and to improve model performance and generalisability, we also examined the impact of using sex and a sex-feature interaction. We trained a binary classification task to distinguish individuals in the Dementia cohort from those in the Population Control. We performed an ID split to generate training and testing subsets in an approximate 80:20 ratio, stratified by cohort. The training data represented 80% of the total data (n = 334 participants) and the testing data represented the remaining 20% (n = 85 participants). During hyperparameter and threshold optimisation, validation sets were produced by splitting the training data (by participant ID) using stratified k-fold cross-validation. See Figure S1 ‘Dementia risk prediction’ in Supplementary Information Section S1 for a visualisation of this evaluation. We used sensitivity, specificity, and the area under the precision-recall curve to measure model performance (for definitions of metrics, please see Supplementary Information Section S3).

#### Risk prediction models

We tested two commonly used classification models: Logistic Regression^74^ and XGBoost.^73^ These models act as effective baselines representing a contrast between linear and non-linear approaches, respectively, with Logistic Regression offering interpretability, while XGBoost provides a more powerful model capable of capturing complex feature interactions. Hyperparameter optimisation was performed using k-fold cross-validation (k = 10), with the model producing the highest mean precision-recall area under the curve score on validation data selected for the final analysis. See Supplementary Information Figure S1 ‘Dementia risk prediction’ in Supplementary Information Section S1 for a visualisation of this evaluation. Supplementary Information Section S5 contains information on the decisions made regarding each step of the classification pipeline. While there was only a slight imbalance in the distribution of individuals from each cohort in the dataset, we once again used this evaluation to explore sample weighting as a strategy to mitigate potential bias towards the overrepresented class, assigning weights inversely proportional to the cohort frequency.

#### Stratification of risk scores for clinical reporting

We extracted probability estimates from the model as a measure of dementia risk. These risk scores were then stratified into three groups (Green, Amber, and Red), denoting low, medium, and high risk of dementia, respectively. These groupings provide a systematic framework to inform clinical decision-making, offering interpretable and data-driven insights. Varying these thresholds allows us to balance sensitivity and specificity with model uncertainty, which can be used to reveal the key difference between individuals the model predicted as being at high and low risk of dementia. This approach was first developed in our group by Capstick *et al*. and has since been used across various research scenarios.^75–77^ Supplementary Information Section S10 provides further information on the process of stratification.

The optimal thresholds used in our final analysis were calculated using k-fold cross-validation (k=5), using the model’s predictions of the validation data after being re-trained with the best hyperparameters on the training data. By choosing 5-fold cross-validation, we limit the amount of model fine-tuning, avoid overfitting to the validation data. See Figure S1 ‘Dementia risk prediction’ in Supplementary Information Section S1 for a visualisation of this evaluation.

## Data Availability

The Withings dataset was provided under a data-sharing agreement for research with Imperial College London and is not publicly available. A subset of the Minder dataset has been made publicly available and can be found on Zenodo at: https://zenodo.org/records/7622128.^78^ A full description of this data subset is published in Nature Scientific Data and can be found here:https://doi.org/10.1038/s41597-023-02519-y.^79^ The extended Minder dataset is available from the corresponding authors upon reasonable request. The Resilient dataset has been made publicly available and can be found on Zenodo at: https://zenodo.org/records/16755408.^80^

## Code Availability

To promote the sharing of resources, the models presented in this paper, as well as all associated code, can be found on GitHub at https://github.com/tmi-lab/Sleep-Age-Dementia. The code for experiments presented in this study will be made available by the corresponding author upon reasonable request.

## Acknowledgements

This study is funded by the UK Dementia Research Institute (UK DRI) Care Research and Technology Centre funded by the Medical Research Council (MRC), Alzheimer’s Research UK, Alzheimer’s Society (grant number: UKDRI–7002), and the UKRI Engineering and Physical Sciences Research Council (EPSRC) Resilient Project (grant number: EP/W031892/1). Infrastructure support for this research was provided by the NIHR Imperial Biomedical Research Centre (BRC) and the UKRI Medical Research Council (MRC). Payam Barnaghi is also funded by the Great Ormond Street Hospital and the Royal Academy of Engineering. The funders were not involved in the study design, data collection, data analysis or writing of the manuscript.

We are incredibly grateful to the participants in our clinical and pilot studies for their contributions to our research and the broader research community.

We would also like to acknowledge support from Surrey and Borders Partnership NHS Foundation Trust and the UK Dementia Research Institute, Care Research and Technology Centre.

## CR&T Group

Acknowledgement list for UK Dementia Research Institute (UK DRI) Care Research & Technology (CR&T) Centre publications using the Minder core data set. The primary contact for this group is

### Leadership and Management

David Sharp (Director), Danielle Wilson (Centre and Research Commercialisation Manager), Sarah Daniels (Hearalth and Social Care Lead), Ramin Nilforooshan (Clinical Lead), David Wingfield (General Practice Lead), Matthew Harrison (Human-Centred Design Lead), Shlomi Haar (Movement Data and Living Lab Lead), Nora Joby (Data Science Lead), Mara Golemme (Scientific Project Manager), Stephanie Lietz (Scientific Project Manager), Margherita Tecilla (Scientific Project Manager).

### Behaviour and Cognition Group

David Sharp (Group Lead), Michael David, Martina Del Giovane, Neil Graham, Magdalena Kolanko, Helen Lai, Lucia M Li, Mark Crook Rumsey, Emma Jane Mallas, Alina-Irina Serban, Eyal Soreq, Abidemi Otaiku, Megan Parkinson, Thomas Parker, Success Fabusoro, Emily Beal, Julian Jeyasingh Jacob, Gaia Frigerio, Anastasia Mirza-Davies, Ethan de Villiers.

### Bioelectronic Systems Group

Timothy Constandinou (Group Lead), Alan Bannon, Danilo Mandic, Ziwei Chen, Charalambos Hadjipanayi, Ghena Hammour, Bryan Hsieh, Amir Nassibi, Adrien Rapeaux, Ian Williams, Maowen Yin, Niro Yogendran.

### Robotics and AI Interfaces Group

Ravi Vaidyanathan (Group Lead), Maria Lima, Ting Su, Melanie Jouaiti, Maitreyee Wairagkar, Carlos Sebastian Castillo, Panipat Wattansiri, Thomas Martineau, Mayue Shi, Tianbo Xu, Alejandro Valdunciel, Reneira Seeamber, Annika Guez, Zehao Liu, Saksham Dhawan, Alina-Irina Serban.

### Translational Machine Intelligence Group

Payam Barnaghi (Group Lead), Nan Fletcher-Lloyd, Samaneh Kouchaki, Alexander Capstick, Chloe Walsh, Louise Rigny, Marirena Bafaloukou, Jin Cui, Yu Chen, Nivedita Bijlani, Iona Biggart, Antigone Fogel, Nathalia Cespedez, Zeinab Ghannam.

### Point of Care Diagnostics Group

Paul Freemont (Group Lead), Michael Crone, Kirsten Jensen, Martin Tran, Thomas Adam, Raphaella Jackson, Alexander Webb, David Wingfield.

### Sleep and Circadian Group

Derk Jan Dijk (Group Lead), Anne Skeldon, Kevin Wells, Ullrich Bartsch, Ciro Della Monica, Kiran KG Ravindran, Victoria L Revell, Hana Hassanin, James Woolley, Iris Wood-Campar, Sarmad Al Gawwam, Aravind Kumar Kamaraj, Marta Pineda Messina.

### Brain and Movement Group

Shlomi Haar (Group Lead), Nathan Steadman, Federico Nardi, Cosima Graef, Alena Kutuzova, Assaf Touboul, Nicolas Calvo Peiro, Jenna Yun, Sean Carr, Uri Rosenblum-Belzer, Emma Burroughs.

### Computational Neurology Group

Gregory Scott (Group Lead), Adela Desowska, Anastasia Gailly de Taurines, Ruxandra Mihai, Nina Moutonnet.

### Human Centred Design Group

Matthew Harrison (Group Lead), Sophie Horrocks, Brian Quan, Victoria Simpson.

### Site Investigators and Key Personnel

#### Surrey and Borders Partnership NHS Foundation Trust

Ramin Nilforooshan (Chief Investigator), Jessica True (Research and Development Manager), Olga Balazikova (Research and Development Manager), Behnam Shariati (Research Doctor), Chloe Walsh (Research Co-ordinator), Nicole Whitethread, Matthew Purnell, Vaiva Zarombaite, Lucy Copps, Olivia Knight, Gaganpreet Bangar, Sumit Dey, Chelsea Mukonda, Jessica Hine, Luke Mallon, Saijal Jhala, Oliver Sargentoni, Amy Alves, Mahan Heydari, Alexandra Cairns (Clinical Monitoring Team).

#### Hammersmith and Fulham Partnership

David Wingfield (Principal Investigator), Monica Morim (Research Nurse/Paramedic), Anesha Patel, Ruby Lyall, Sanara Raza, Success Fabusoro, Gaia Frigerio, Maria Rasulo, Catalina Chavarro Novoa, Martynas Stonkus, Prital Patel, Zara Prem (Clinical Studies), Naomi Hassim, Pippa Kirby (Research Allied Healthcare Professionals) John Patterson(London Borough of Hammersmith and Fulham Support: Assistive Technology), Mike Law (Business Development), Andy Kenny (Social Services).

#### Minder Digitally Enabled Care for Dementia (MinderCare)

David Sharp (Principal Investigator), James Bird (Co-Investigator), Sarah Pearse (Co-Investigator), Joanna James, Janibo Amade Cassimo, Aglaja Dar, Pandora Wright, Lucia Li, Anastasia Mirza-Davies, Julian Jeyasingh Jacobs, Zinca Zecevic, Sarah Daniels, David Wingfield (Clinical Team), Success Fabusoro, Gaia Frigerio, Maria Rasulo, Catalina Chavarro Novoa (Research Technicians).

## Author Contributions

NFL: Data Curation, Conceptualisation, Methodology, Software, Data Processing, Investigation, Formal Analysis, Pilot Analysis, Writing—Original Draft, Review and Editing, Visualisation; NCG: Data Curation, Methodology, Software, Data Processing, Investigation, Writing-Original Draft, Review and Editing; ACapstick: Data Curation, Conceptualisation, Methodology, Software, Data Processing, Investigation; AF, MB: Writing-Original Draft, Review and Editing; MH, ACairns: Data Curation; CW, JT: Data Curation, Project Administration; CR&T: Data Curation (Minder data); BS: Data Curation, Pilot Analysis; RN: Clinical Study Lead, Conceptualisation, Data Curation, Pilot Analysis, Review and Editing, Supervision; and PB: Data Curation, Conceptualisation, Methodology, Investigation, Formal Analysis, Pilot Analysis, Review and Editing, Supervision, Funding Acquisition.

