## Supplementary Information for "Real-world deployment of remote sleep monitoring technologies reveals distinct patterns associated with cognitive decline"

### S1 Study workflow and experimental setup

As described in the Methodology Section, we use a two-stage experimental pipeline to predict dementia risk using sleep age estimated from nocturnal activity and physiology data collected using under-the-mattress sleep sensors (see Figure S1).

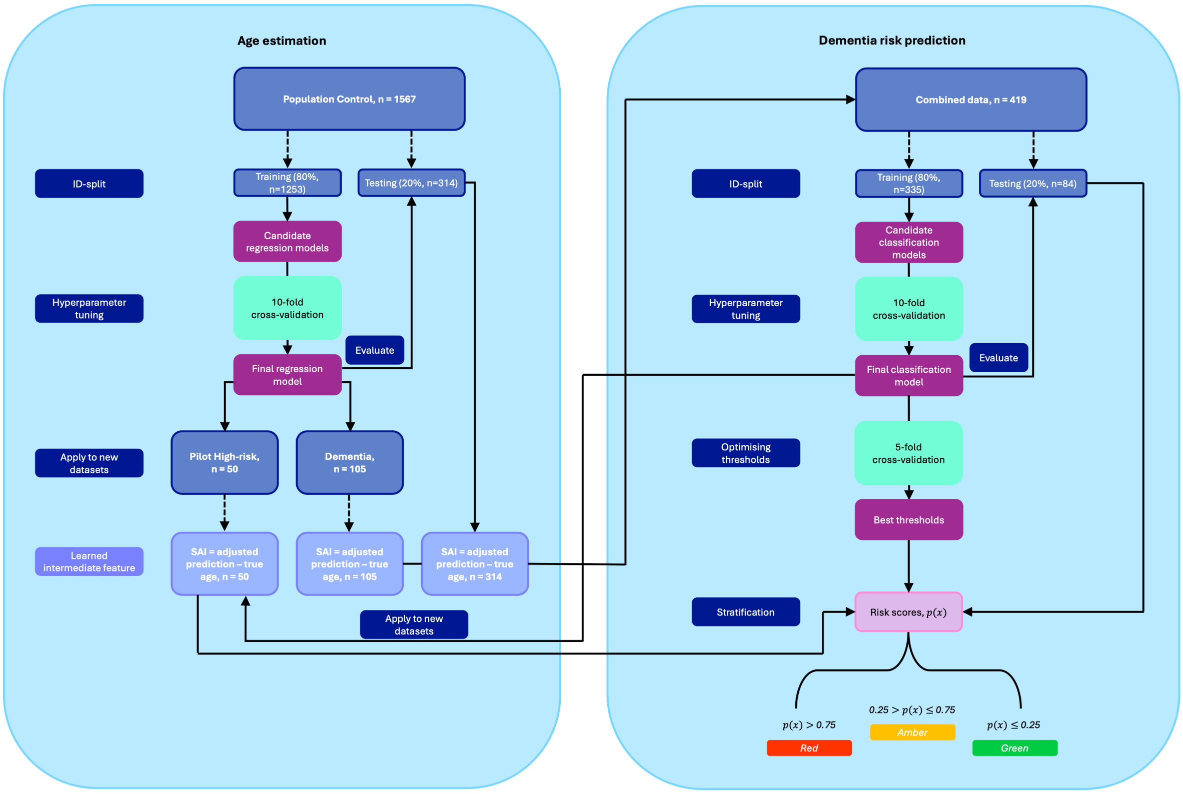


**Figure S1:** Overview of the study workflow and experimental setup. This figure illustrates the data used at each stage of the pipeline, the evaluation procedures, and the final outputs of the study.

### S2 Demographics and clinical characteristics

In this section, we provide information on the demographics and clinical characteristics of the three cohorts datasets presented in this work. Table S1 reports the demographics of the three datasets before and after data processing. Table S2 reports the primary diagnoses for the Dementia cohort participants, as well as the mean and standard deviation of the Mini-Mental State Examination (MMSE) used to assess the severity of cognitive impairment. Table S3 shows the corresponding severity classifications. Tables S4 and S5 report the top five most common diagnoses and medications among the participants in the Pilot High-risk cohort, and Table S6 reports the Addenbrooke's Cognitive Examination-III (ACE-III) scores and respective severity classifications used to assess the severity of cognitive impairment. Tables S7 and S8 report the Patient Health Questionnaire-9 (PHQ-9) and Generalised Anxiety Disorder Questionnaire-7 (GAD-7) scores, as well as the respective severity classifications used to assess the severity of depression and anxiety.

### S3 Evaluation metrics

Age is associated with both an increased risk of dementia and significant changes in sleep architecture, which may contribute to the development or progression of dementia. In this work, we have proposed a two-stage experimental pipeline to predict dementia risk from the Sleep Age Index (SAI) estimated from remote sleep monitoring data. At each stage of the pipeline (age estimation and dementia risk prediction), we assessed the performance of our models to ensure both the usefulness and reliability of our models.

**Table S1:** Cohort demographics.

| Cohort | Dataset | Sample size, n | Sex, (Female, Male) | Age range |
|---|---|---|---|---|
| **Population Control** | Original | 1,080,081 | F=1,034, M=4,540 | 19-99 |
| **Population Control** | Processed | 15,229 | F=326, M=1,241 | 50-98 |
| **Dementia** | Original | 116,862 | F=82, M=78 | 51-100 |
| **Dementia** | Processed | 3,140 | F=40, M=65 | 52-92 |
| **Pilot High-risk** | Original | 14,412 | F=44, M=30 | 72-98 |
| **Pilot High-risk** | Processed | 470 | F=29, M=21 | 72-92 |

**Table S2:** Primary diagnosis and MMSE scores in Dementia cohort. Scores given to 2.d.p. *MCI* mild cognitive impairment, *MMSE* Mini-Mental State Examination.

| Primary diagnosis | % of cohort | MMSE score |
|---|---|---|
| **Alzheimer's** | 40.0 | 19.6 (±6.78) |
| **Frontotemporal dementia** | 2.50 | 22.3 (±8.47) |
| **Mixed dementia** | 11.9 | 18.9 (±7.80) |
| **Vascular dementia** | 6.88 | 19.4 (±7.82) |
| **MCI** | 2.50 | 25.1 (±3.90) |
| **Other** | 11.3 | 25.8 (±3.96) |
| **Not known** | 25.0 | 21.6(±7.74) |

**Table S3:** MMSE scores and severity classifications in Dementia cohort. *MMSE* Mini-Mental State Examination.

| Impairment Severity | MMSE Score |
|---|---|
| **Normal Cognition** | 27 - 30 |
| **Mild** | 21 - 26 |
| **Moderate** | 15 - 20 |
| **Moderately Severe** | 10 - 14 |
| **Severe** | 0 - 9 |

**Table S4:** Top five most common diagnoses of participants in Pilot High-risk cohort.

| Diagnosis | % of cohort |
|---|---|
| **Essential Hypertension** | 77.4 |
| **Osteoarthritis** | 45.3 |
| **Cataracts** | 39.6 |
| **Asthma** | 24.5 |
| **Diverticulosis** | 24.5 |

*Note.* Medications were not recorded for 2 study participants.

**Table S5:** Top five most common medications of participants in Pilot High-risk cohort.

| Medication | % of cohort |
|---|---|
| **Other** | 76.4 |
| **Atorvastatin** | 38.2 |
| **Lansoprazole** | 32.7 |
| **Bisoprolol** | 23.6 |
| **Ramipril** | 20.0 |

*Note.* Medications were not recorded for 2 study participants.

**Table S6:** ACE-III scores and severity classifications of study participants in Pilot High-risk cohort on entry into clinical study. *MCI* mild cognitive impairment, *ACE-III* Addenbrooke's Cognitive Examination-III.

| Severity | No. of Participants | % of Cohort |
|---|---|---|
| **Normal Cognition (ACE-III $\geq$ 88)** | 27 | 49.1 |
| **Possible MCI (84 $\leq$ ACE-III $\geq$ 87)** | 13 | 23.6 |
| **Suspected Dementia (76 $\leq$ ACE-III $\geq$ 83)** | 8 | 14.6 |
| **Likely Dementia (ACE-III $<$ 76)** | 7 | 12.7 |

**Table S7:** PHQ-9 scores and severity classifications of study participants in Pilot High-risk cohort on entry into clinical study. *PHQ-9* Patient Health Questionnaire-9.

| Severity | No. of Participants | % of Cohort |
|---|---|---|
| **Minimal or No Depression (PHQ-9 $<$ 5)** | 23 | 41.8 |
| **Mild Depression (5 $\leq$ PHQ-9 $\geq$ 9)** | 24 | 43.6 |
| **Moderate Depression (10 $\leq$ PHQ-9 $\geq$ 14)** | 5 | 9.10 |
| **Moderately Severe Depression (15 $\leq$ PHQ-9 $\geq$ 19)** | 2 | 3.64 |
| **Severe Depression (PHQ-9 $\geq$ 20)** | 1 | 1.82 |

#### S3.1 Age estimation

In the age estimation stage of our pipeline, we measured the performance of the proposed regression models using three evaluation metrics: mean absolute error (MAE), mean squared error (MSE), and balanced mean absolute error (BMAE).

MAE assesses how far off the predicted value is from the actual value, measuring the average size of the errors, without considering their direction (under or over). MSE measures the average squared difference between the predicted and actual values, penalising large errors more heavily, making this metric more sensitive to outliers. Low MAE and MSE values indicate that the model's predictions are close to the actual values, while a low MSE also indicates that the model rarely makes large errors.

$$MAE = \frac{1}{n}\sum_{i=1}^{n}|y_i - \hat{y}_i| \tag{1}$$

$$MSE = \frac{1}{n}\sum_{i=1}^{n}(y_i - \hat{y}_i)^2 \tag{2}$$

where $y_i$, $\hat{y}_i$, and $n$ refer to the actual value, predicted value, and number of observations, respectively.

As older age groups are less represented in the Population Control cohort, we use the BMAE to ensure that each age group contributes equally to the final error. This avoids letting majority groups dominate the error calculation, making the measure of model performance fairer and more uniform. A low MAE indicates that the model performs well across all groups, regardless of the number of data points in each age group.

$$BMAE = \frac{1}{G}\sum_{g=1}^{G}\left(\frac{1}{n_g}\sum_{i\in g}|y_i - \hat{y}_i|\right) \tag{3}$$

**Table S8:** GAD-7 scores and severity classifications of study participants in Pilot High-risk cohort on entry into clinical study. *GAD-7* Generalised Anxiety Disorder Questionnaire-7.

| Severity | No. of Participants | % of Cohort |
|---|---|---|
| **Minimal or No Anxiety (GAD-7 $<$ 5)** | 44 | 80.0 |
| **Mild Anxiety (5 $\leq$ GAD-7 $\geq$ 9)** | 7 | 12.7 |
| **Moderate Anxiety (10 $\leq$ GAD-7 $\geq$ 14)** | 2 | 3.64 |
| **Severe Anxiety (GAD-7 $\geq$ 14)** | 2 | 3.64 |

where $G$ refers to the number of groups, $n_g$ refers to the number of samples in group $g$, and $\sum_{i \in g}$ means sample $i$ belongs to group $g$, respectively.

The combined use of these metrics helps provide a comprehensive picture of the model's ability to estimate age.

#### S3.2 Dementia risk prediction

In the dementia risk prediction stage of our pipeline, we measured the performance of the proposed classification models using three evaluation metrics: sensitivity, specificity, and area under the precision-recall curve (AUC Precision-Recall).

Sensitivity (also known as recall) measures the proportion of true positive instances correctly identified by the model, assessing how well the model identifies individuals from the Dementia cohort. In contrast, specificity measures how well the model identifies individuals from the Population Control cohort. A high sensitivity indicates that the model is effective at identifying individuals with dementia, while a high specificity indicates that the model is effective at identifying individuals within the Population Control cohort.

$$Sensitivity = \frac{\text{TP}}{\text{TP} + \text{FN}} \tag{4}$$

$$Specificity = \frac{\text{TN}}{\text{TN} + \text{FP}} \tag{5}$$

where TP, TN, FP, and FN refer to True Positives, True Negatives, False Positives, and False Negatives, respectively.

Finally, we use the AUC Precision-Recall to optimise the hyperparameters of our classification model. The AUC Precision-Recall is the area under the curve created by plotting the precision of a model against its recall, measuring how well the model identifies the positive class across all possible thresholds. A high AUC Precision-Recall indicates that the model is effective at correctly identifying individuals with dementia while maintaining a low rate of false positives (high precision). Precision is the proportion of correct positive predictions. It is a measure of how confident the model is that the positive prediction is actually correct. A high precision indicates that when the model identifies an individual as having dementia, it is likely correct.

$$Precision = \frac{\text{TP}}{\text{TP} + \text{FP}} \tag{6}$$

The combined use of these metrics helps provide a comprehensive picture of the model's ability to identify individuals with dementia.

### S4 Results of preliminary statistical analyses

In this section, we report the results of the preliminary analyses conducted on the processed Population Control dataset. Figure S2 visualises the comparison of the statistics (mean, median, minimum, maximum, standard deviation, and interquartile range) derived for each of 16 sleep features ('Time to Bed (angles)', 'Time to Rise (angles)', 'Time in Bed (s)', 'Time Out of Bed (s)', 'Bed Time Period (s)' (total time in bed), 'Rapid eye movement (REM) Duration (s)', 'Light Duration (s)', 'Deep Duration (s)', 'Awake Duration (s)', 'Time Snoring (s)', 'Heart rate (HR) Average (beats per minute or BPM)', 'HR Minimum (BPM)', 'HR Maximum (BPM)', 'Respiratory rate (RR) Average (breaths per minute or BPM)', 'RR Minimum (BPM)', 'RR Maximum (BPM)') between four age bands (50-59, 60-69, 70-79, 80+), with standard significance reporting is used to denote differences between age groups ($p < 0.0001$ denoted as ***, $p < 0.001$ denoted as **, $p < 0.05$ denoted as *, and $p < 0.1$ denoted as $\cdot$).

### S5 Machine learning pipeline evaluated

For each stage of this work, we evaluated several combinations of models, as well as the data used as the initial input to our two-stage pipeline. In this section, we describe the different techniques we experimented with before arriving at our final pipeline.

#### S5.1 Data

This section presents the number of unique participants and total samples by each of the four age bands (50-59, 60-69, 70-79, and 80+), across n-day rolling window lengths. We compared three different window lengths. Figure S3 shows bar charts comparing n = 30 (Figure S3 (a)), n = 60 (Figure S3 (b)), and n = 90 (Figure S3 (c)) window lengths.

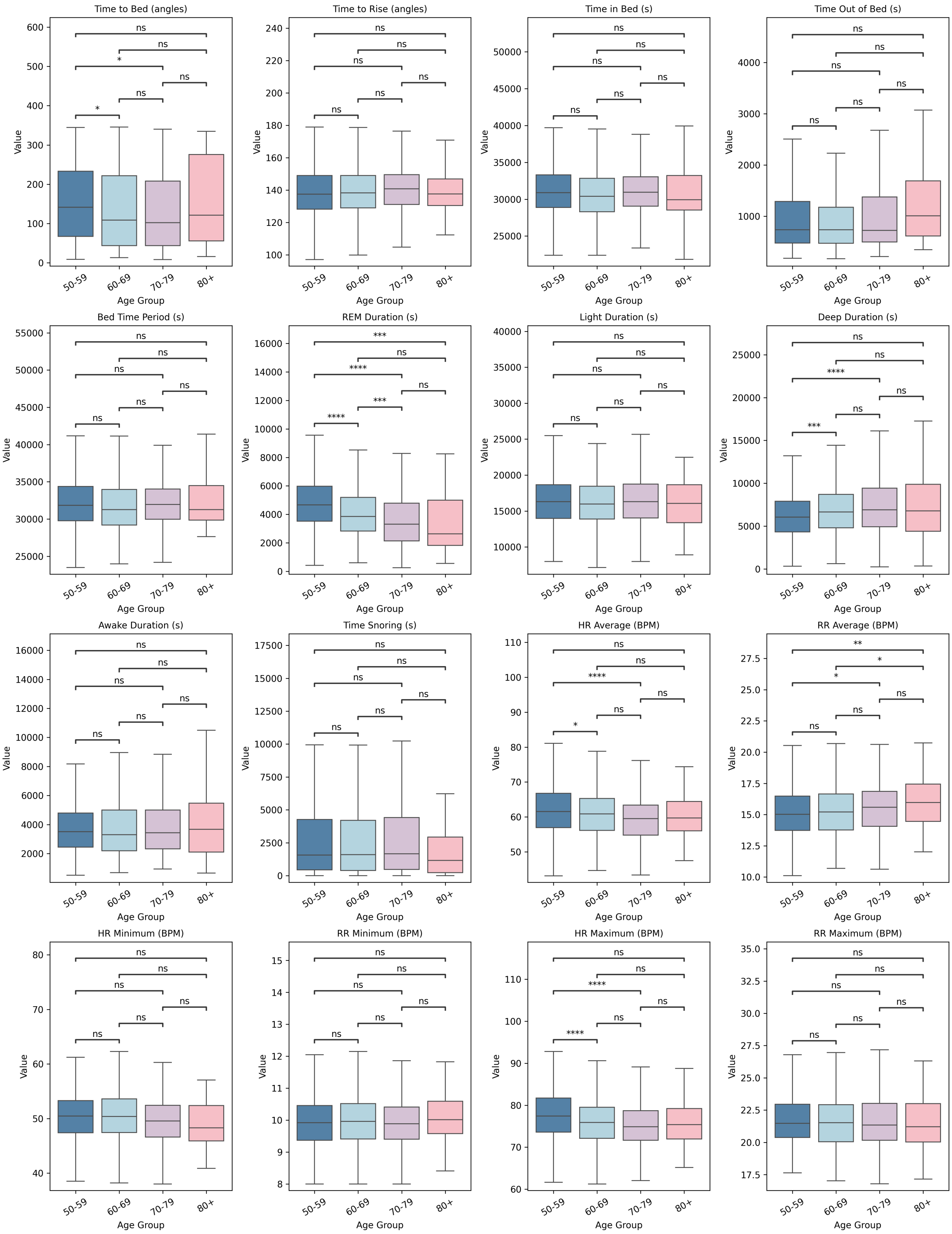


**Figure S2:** Boxplots of statistical measures (a–f) of sleep features compared across age groups, with standard significance reporting ($p < 0.0001$ denoted as ***, $p < 0.001$ denoted as **, $p < 0.05$ denoted as *, and $p < 0.1$ denoted as ·). **(a)** Mean for each feature.

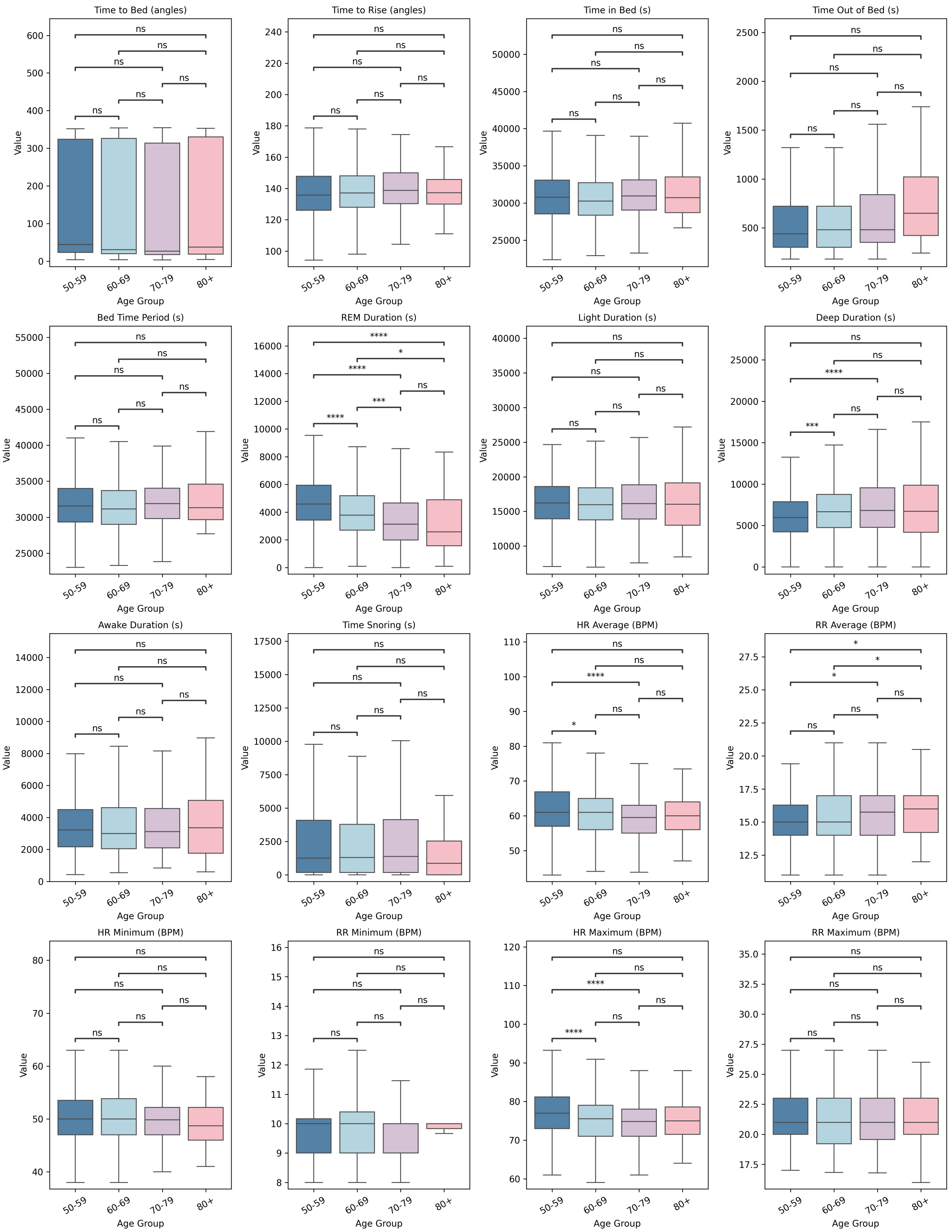


**(b)** Median for each feature.

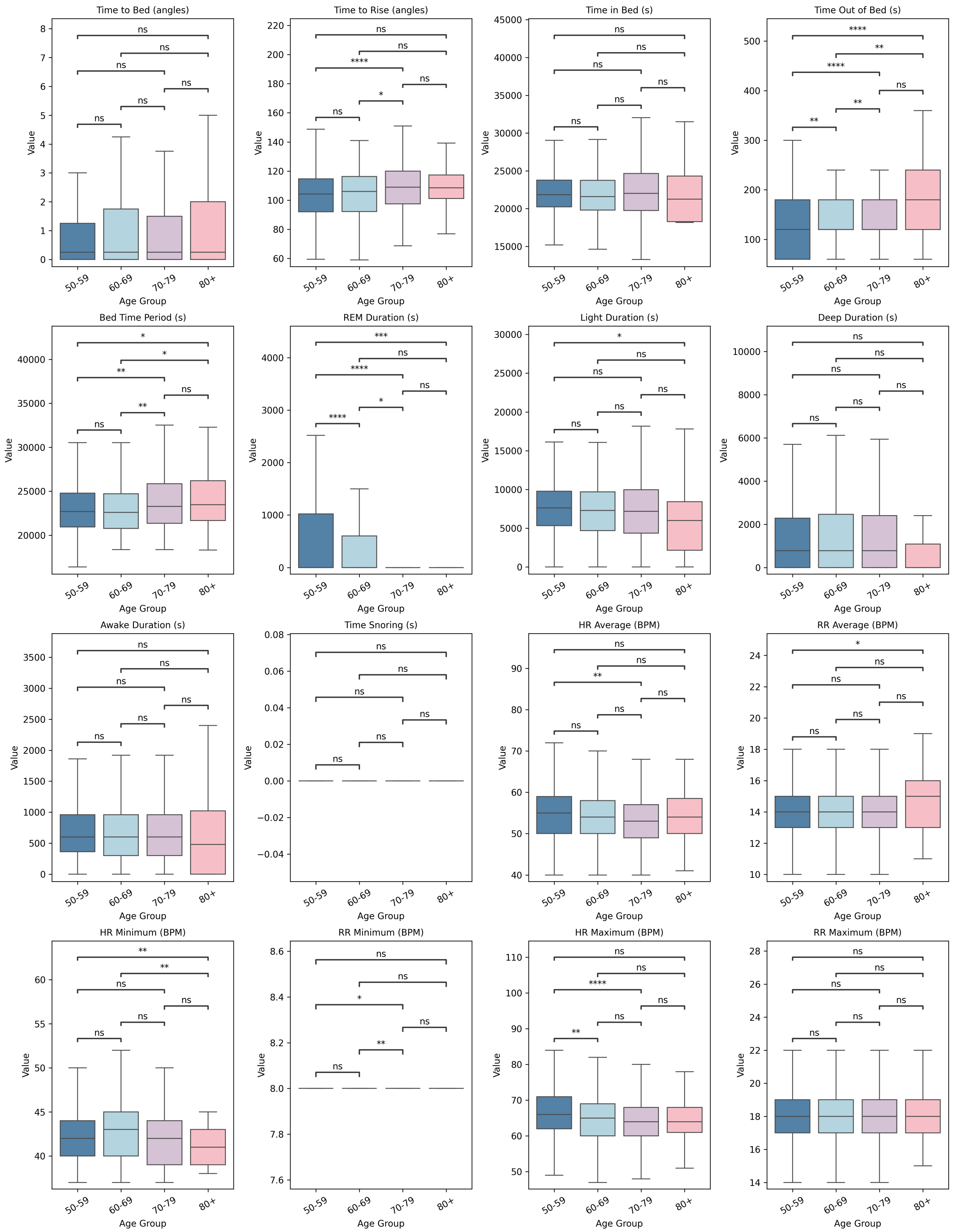


**(c)** Minimum for each feature.

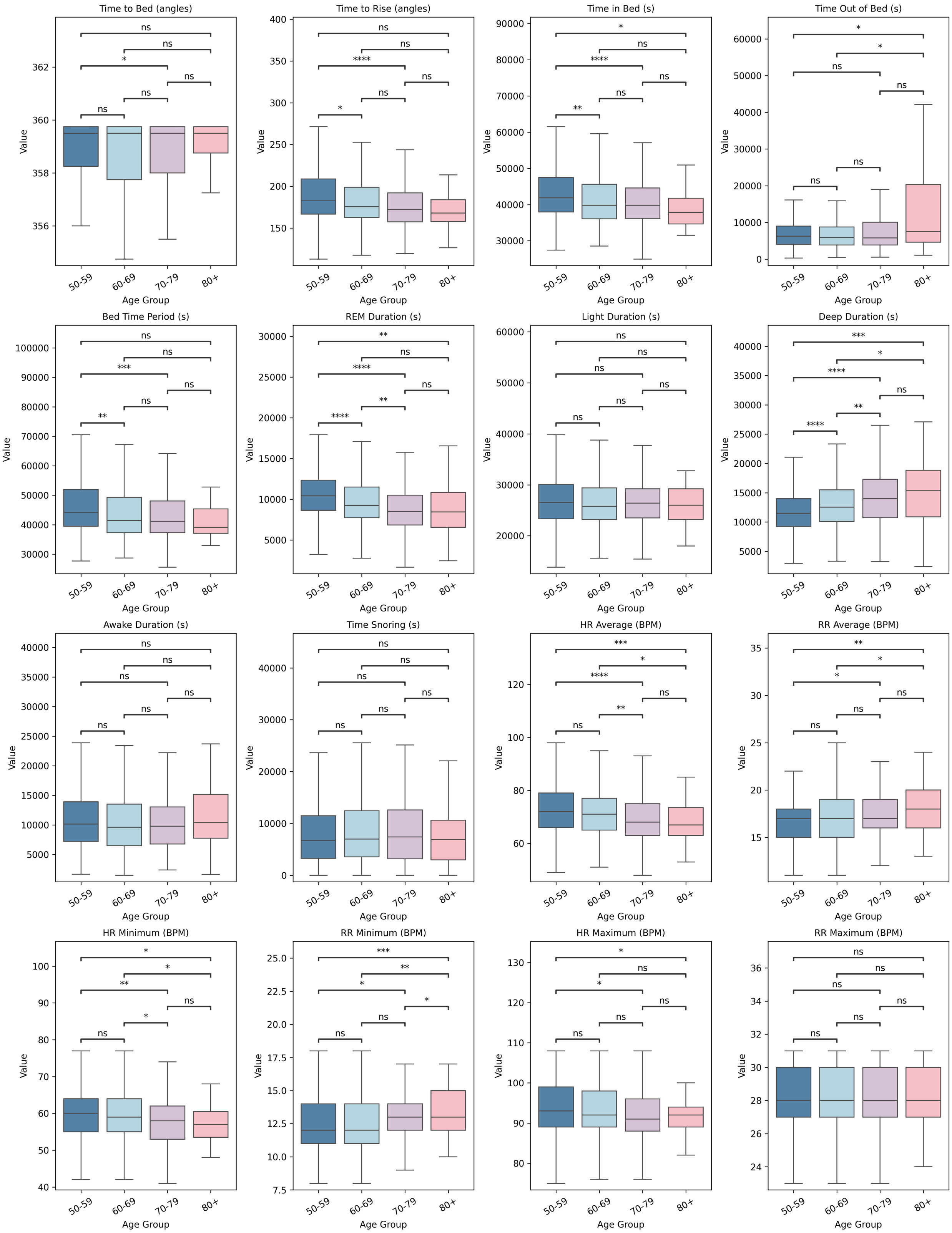


**(d)** Maximum for each feature.

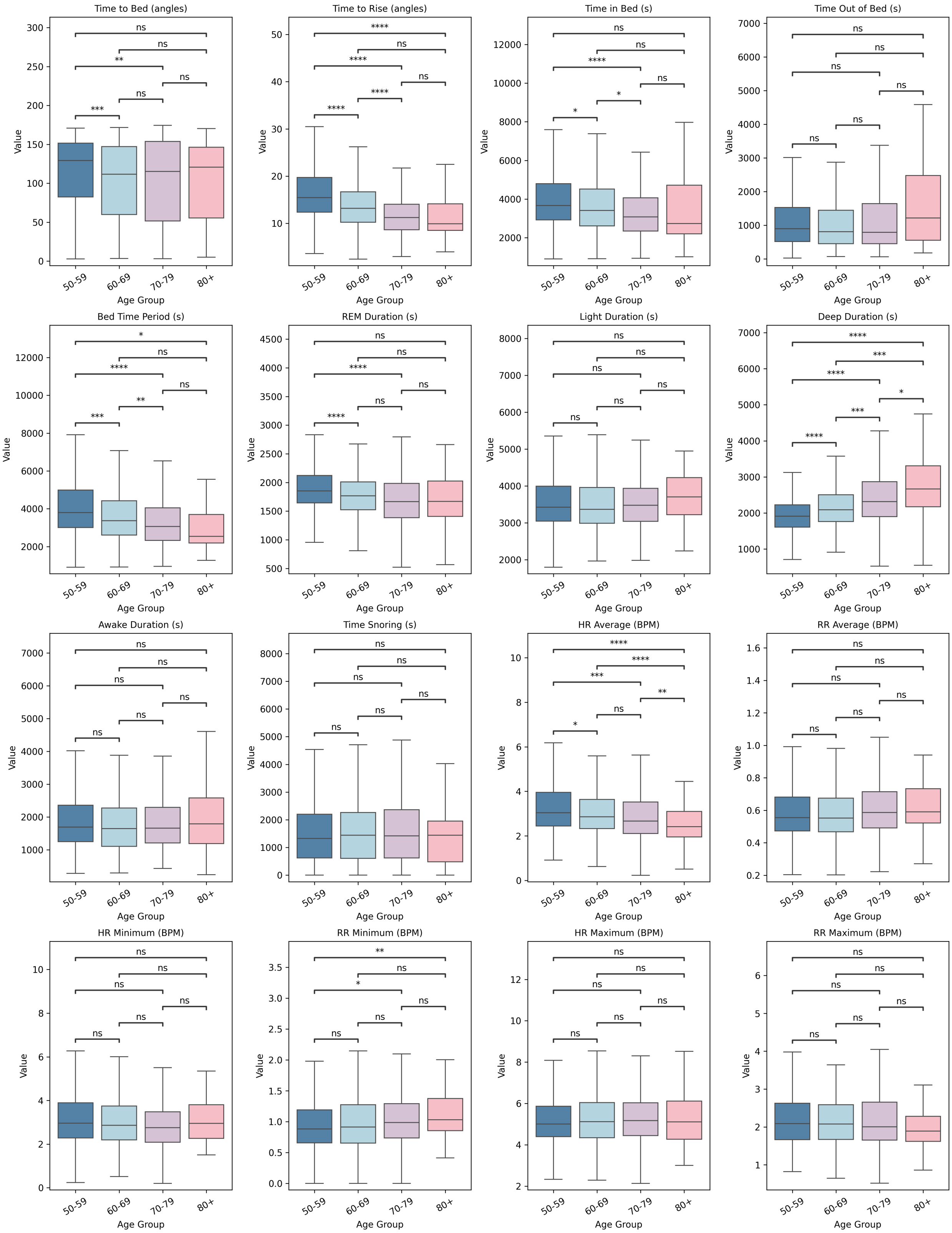


**(e)** Standard deviation for each feature.

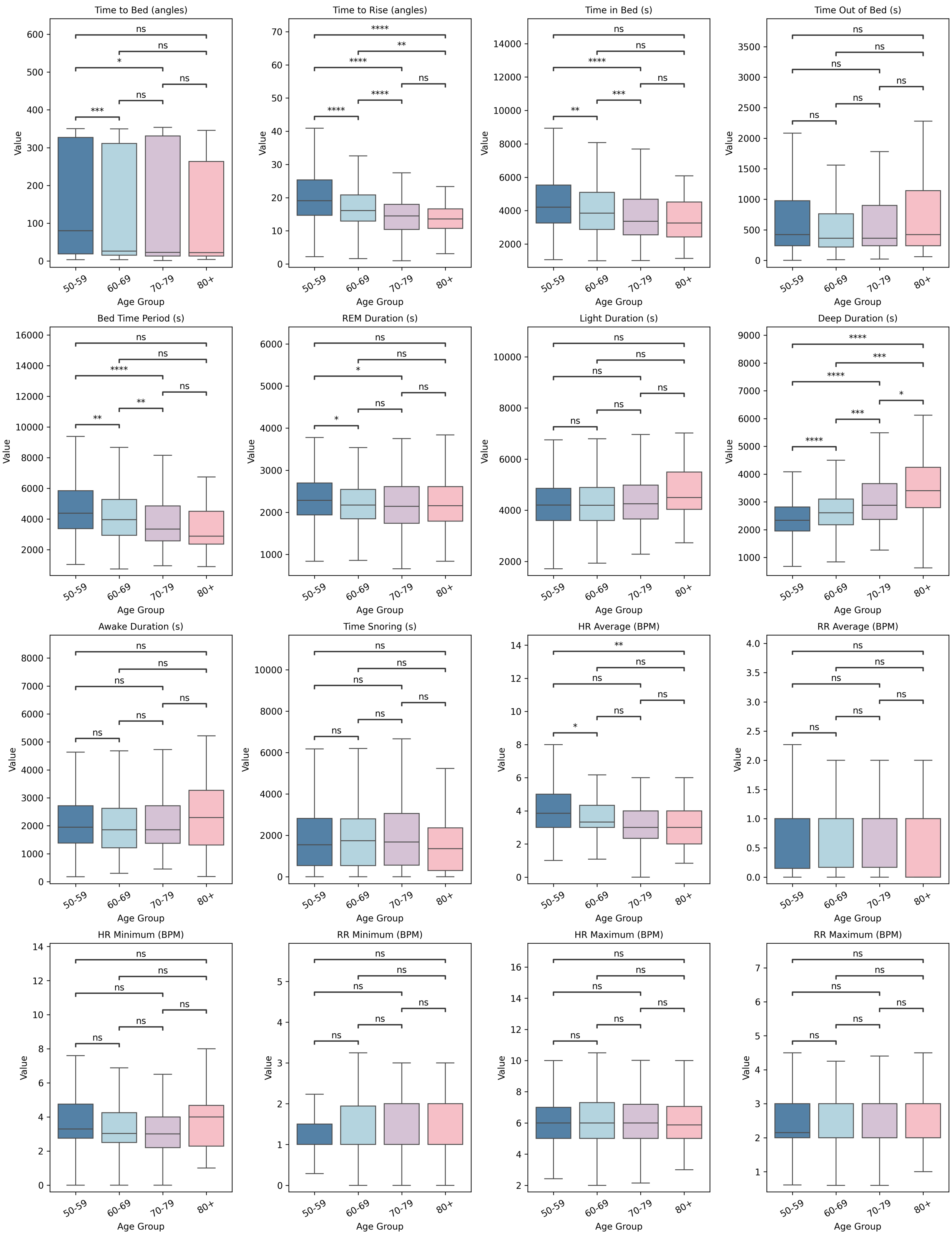


**(f)** Interquartile range for each feature.

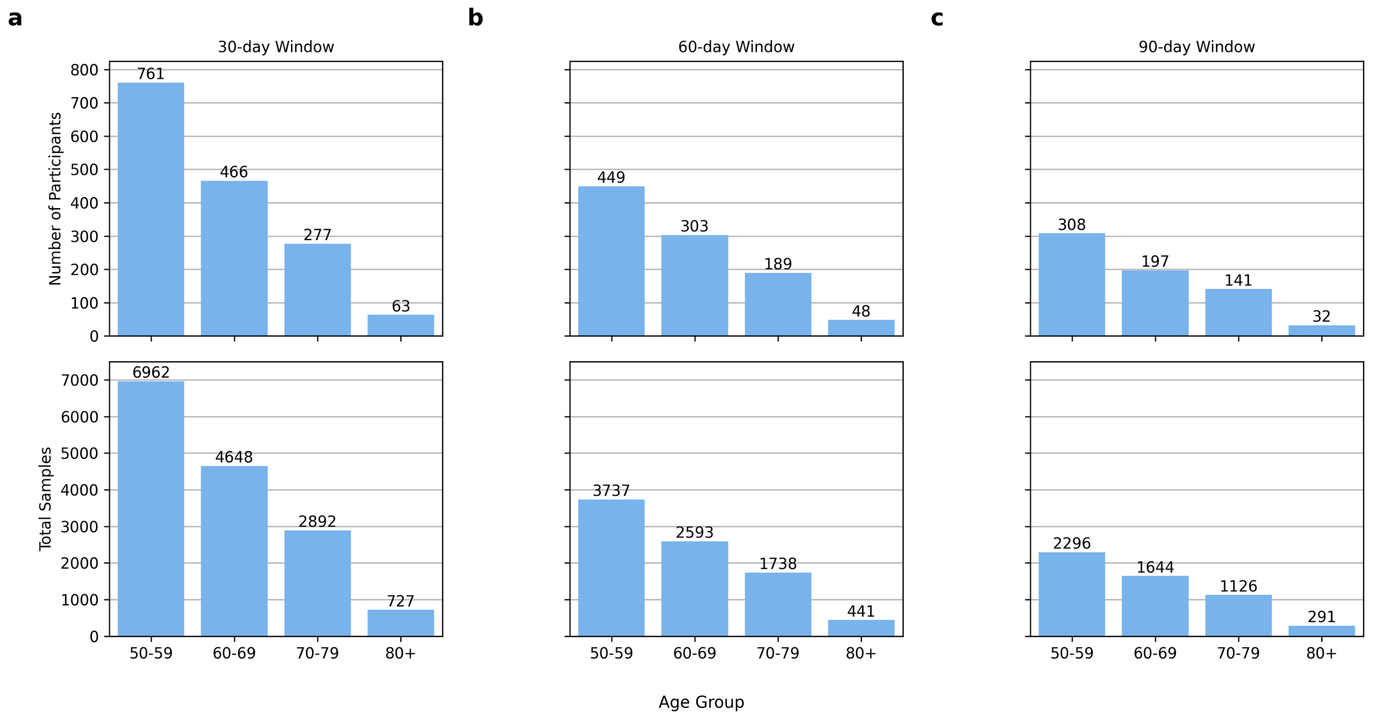


**Figure S3:** Number of unique participants (top row) and total samples (bottom row) for each age group across 30, 60 and 90-day windows.

### S5.2 Machine learning models

We evaluated a range of models at both stages of our pipeline. These models were run using Python and implemented using Sci-kit Learn.[65] The final model was implemented using the best-performing hyperparameters found across a 10-fold cross-validation.

**Regression models** We evaluated three commonly used regression models: Elastic Net, Huber, and Extreme Gradient Boosting (XGBoost).

For all models, we used Bayesian optimisation to determine the best values of the following hyperparameters:

- Elastic Net: With overall regularisation strength between [1e-5, 1e+3], and the balance between L1 and L2 penalties in [0.0, 1.0].
- Huber: With L2 regularisation, with a value between [1e-5, 1e-1], and a loss threshold between [1.0, 10.0].
- Extreme Gradient Boosting (XGBoost): with a learning rate between [0.01, 0.1], number of estimators between [50, 500]. max tree depth of [5, 10], sub-sample ratio between [0.5, 1.0], and L1 regularisation between [1.0, 10.0].

**Classification models** We evaluated two commonly used classification models: Logistic Regression (LR), and Extreme Gradient Boosting (XGBoost).

For both models, we used Bayesian optimisation to determine the best values of the following hyperparameters:

- Logistic Regression (LR): With L2 regularisation, with a value between [0.001, 1], determined by hyperparameter optimisation.
- Extreme Gradient Boosting (XGBoost): with a learning rate between [0.01, 0.1], number of estimators between [50, 500]. max tree depth of [5, 10], sub-sample ratio between [0.5, 1.0], and L1 regularisation between [1.0, 10.0].

**Table S9:** Mean (95% CI) % of MAE, MSE, and BMAE of all age estimation models, with and without sample weighting, on statistics derived from 30, 60, and 90-day arrays of data, across 10-fold cross-validation. *MAE* Mean Absolute Error, *MSE* Mean Squared Error, *BMAE* Balanced Mean Absolute Error.

| Model | No. of Days | Sample Weighting | MAE | MSE | BMAE |
|---|---|---|---|---|---|
| Elastic Net | 30 | w/o | 6.36 (6.10 - 6.62) | 76.8 (57.6 - 110) | 7.14 (6.73 - 7.62) |
| Elastic Net | 30 | w/ | 6.61 (6.34 - 6.89) | 71.8 (61.8 - 87.2) | 7.01 (6.66 - 7.37) |
| Elastic Net | 60 | w/o | 6.27 (5.89 - 6.63) | 63.8 (53.6 - 74.6) | 6.81 (6.32 - 7.29) |
| Elastic Net | 60 | w/ | 6.46 (6.07 - 6.85) | 68.5 (57.4 - 81.8) | 6.74 (6.25 - 7.22) |
| Elastic Net | 90 | w/o | 6.22 (5.74 - 6.75) | 60.4 (51.1 - 71.6) | 6.83 (6.26 - 7.47) |
| Elastic Net | 90 | w/ | 6.48 (6.01 - 6.98) | 63.6 (54.5 - 73.3) | 6.88 (6.27 - 7.51) |
| Huber | 30 | w/o | 6.36 (6.10 - 6.61) | 74.4 (57.7 - 102) | 7.12 (6.72 - 7.58) |
| Huber | 30 | w/ | 6.60 (6.32 - 6.87) | 76.2 (63.0 - 98.1) | 7.04 (6.67 - 7.44) |
| Huber | 60 | w/o | 6.31 (5.92 - 6.68) | 66.1 (54.4 - 79.7) | 6.84 (6.34 - 7.32) |
| Huber | 60 | w/ | 6.51 (6.10 - 6.91) | 70.1 (58.3 - 84.1) | 6.78 (6.27 - 7.27) |
| Huber | 90 | w/o | 6.33 (5.82 - 6.89) | 62.6 (52.8 - 74.2) | 6.92 (6.34 - 7.57) |
| Huber | 90 | w/ | 6.60 (6.06 - 7.16) | 67.5 (56.8 - 79.0) | 7.01 (6.36 - 7.70) |
| XGBoost | 30 | w/o | 5.97 (5.73 - 6.19) | 56.0 (52.0 - 59.7) | 6.59 (6.29 - 6.87) |
| XGBoost | 30 | w/ | 6.07 (5.87 - 6.29) | 57.3 (54.4 - 60.2) | 6.56 (6.34 - 6.80) |
| XGBoost | 60 | w/o | 6.13 (5.68 - 6.54) | 58.7 (50.3 - 66.3) | 6.63 (6.08 - 7.12) |
| XGBoost | 60 | w/ | 6.15 (5.71 - 6.56) | 59.1 (51.3 - 66.4) | 6.55 (6.05 - 7.01) |
| XGBoost | 90 | w/o | 5.89 (5.42 - 6.39) | 55.8 (47.5 - 64.6) | 6.45 (5.92 - 6.98) |
| XGBoost | 90 | w/ | 6.01 (5.55 - 6.50) | 57.4 (48.6 - 67.0) | 6.51 (5.99 - 7.03) |

### S6 Evaluation of age estimation models

Table S9 shows the results of the 10-fold cross-validation on all the models tested on the Population Control training data.

Table S10 presents the model's performance for each sex. We report the mean and standard deviation of the mean absolute error (MAE) for both the Population Control and Dementia cohorts.

**Table S10:** Mean and standard deviation (Std) of MAE for each sex across both the Population Control (PC) and Dementia (D) cohorts. All values are reported to 3.s.f.

| Sex | Cohort (PC \| D) | Mean | Std |
|---|---|---|---|
| Female | PC | 6.03 | 4.52 |
| | D | 14.8 | 7.53 |
| Male | PC | 5.39 | 4.21 |
| | D | 15.4 | 7.13 |

We calculate Cohen's d effect size within each sex. For both the Population Control test set (Cohen's d = 0.15 (3.s.f.)) and the Dementia cohort (Cohen's d = -0.08 (3.s.f.)), there was a small effect size, showing that age prediction performed similarly between sexes.

### S7 Interpretation and analysis of age estimation in the Dementia cohort

In this section, we show the results of additional interpretability and statistical analysis on the outputs of our age estimation model on the Dementia cohort.

#### S7.1 Explainability of age estimation

Figure S4 shows the key sleep metrics that contributed to age prediction for the Dementia cohort. All but one of the top 10 features is the same as seen for the held-out Population Control test set. The magnitude and direction of these effects also remain constant.

#### S7.2 Statistical analysis of SAI across different levels of cognitive impairment

Here, we report the results of a statistical analysis of SAI in the Dementia cohort. Figure S5 illustrates the comparison of mean SAI across the five different severity classifications of cognitive impairment (Normal, Mild, Moderate, Moderately Severe, and Severe), with standard significance reporting is used to denote differences between age groups ($p < 0.0001$ denoted as ***, $p < 0.001$ denoted as **, $p < 0.05$ denoted as *, and $p < 0.1$ denoted as $\cdot$).

### S8 Evaluation of dementia risk prediction models

Table S11 shows the results of the 10-fold cross-validation on all the models tested on on the combined cohort (Population Control and Dementia) SAI.

### S9 Reliability and calibration

In this section, we present the results of the reliability and calibration analysis conducted on the dementia risk prediction model. Figure S6 shows a multi-faceted summary of model performance. The plots on the left-hand side of the figure are reliability plots, showing how well the predicted probabilities of the model align with the true labels (Population Control cohort and the Dementia cohort). These plots show that our dementia risk scores can be considered reasonably reliable when considering both positive and negative predictions together. This is indicated by the small reliability gaps observed in the majority of the plots (a) and the similarity between the average accuracy and average confidence in plot (c). When considering only positive predictions as in plot (b) showing how well the predicted probabilities of the best model correspond to the true likelihood of the positive case, we observe that the model was at times somewhat overconfident (around a mean predicted probability of 0.5) and at others (a mean predicted probability of 0.1 and 0.9) slightly less confident. Finally, plot (d) shows the distribution of the predicted probabilities made by the model on each class, and here we observe that the model was more uncertain about samples taken from individuals in the Population Control cohort, occasionally predicting them to have dementia based on the application of binary thresholds. These results were a key motivation for the use of stratification to ensure that the number of false positives could be controlled.

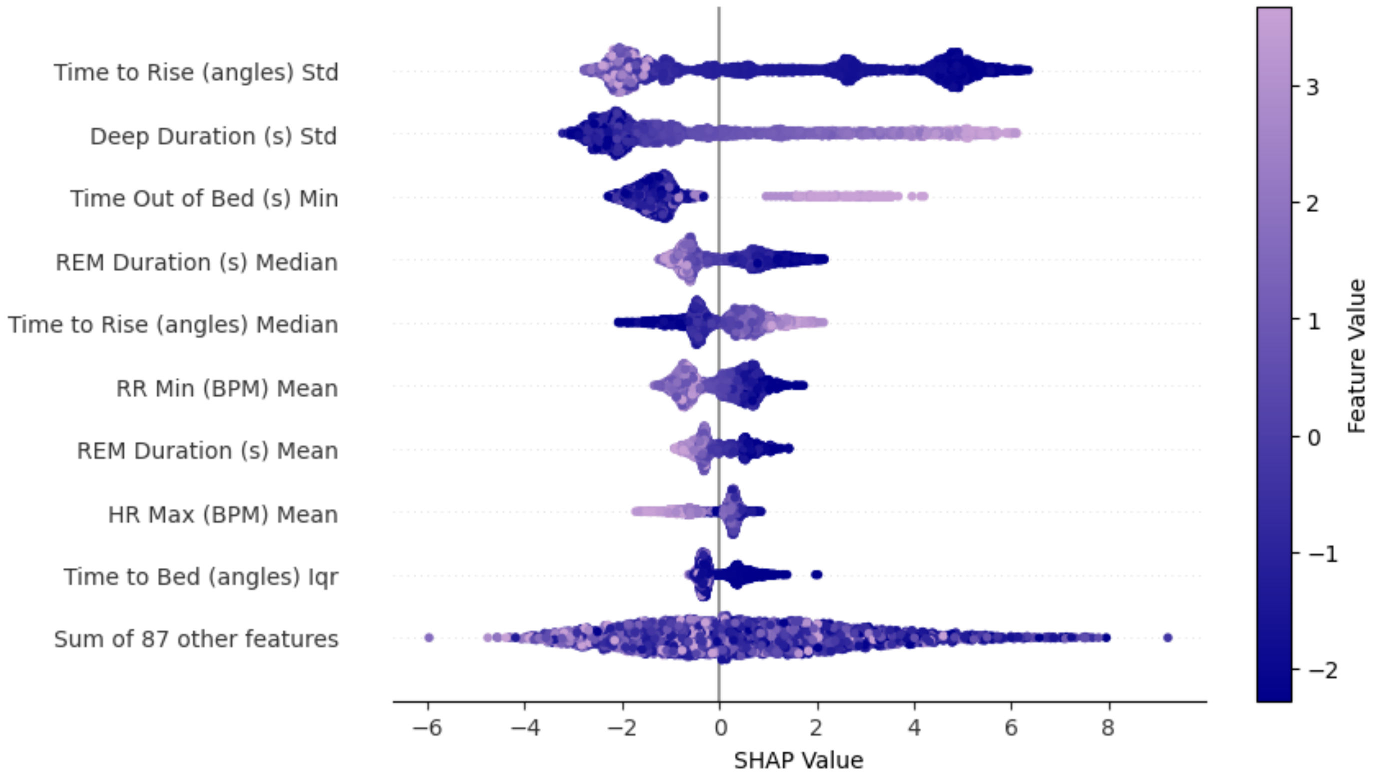


**Figure S4:** SHAP values. The feature importance for the top 10 most important features, as calculated on the Dementia cohort using SHAP. The corresponding feature values are coloured according to the value of that feature (high values shown in lilac and low values in dark blue), whilst the position along the x-axis represents the contribution that particular value made to the prediction.

**Table S11:** Mean (95% CI) % of sensitivity, specificity, and area under the precision-recall curve of all dementia risk prediction models, with and without sample weighting, based on single, across 10-fold cross-validation.

| Model | Sex and Interaction | Sample Weighting | Sensitivity | Specificity | AUC Precision-Recall |
|---|---|---|---|---|---|
| LR | w/o | w/o | 66.0 (57.9 - 73.5) | 71.1 (66.4 - 75.5) | 77.0 (68.7 - 84.7) |
| LR | w/ | w/o | 68.8 (61.2 - 75.9) | 71.5 (69.0 - 73.8) | 78.8 (71.7 - 85.3) |
| LR | w/o | w/ | 65.4 (56.8 - 73.3) | 72.1 (68.3 - 75.7) | 77.0 (68.7 - 84.7) |
| LR | w/ | w/ | 68.7 (61.0 - 75.8) | 72.0 (69.7 - 74.2) | 78.8 (71.7 - 85.3) |
| XGBoost | w/o | w/o | 56.3 (47.7 - 64.2) | 82.7 (79.7 - 85.9) | 76.7 (68.7 - 84.3) |
| XGBoost | w/ | w/o | 58.9 (51.2 - 66.5) | 77.9 (74.7 - 81.2) | 77.2 (70.1 - 83.8) |
| XGBoost | w/o | w/ | 56.5 (47.6 - 64.7) | 83.3 (80.8 - 85.6) | 76.7 (68.7 - 84.3) |
| XGBoost | w/ | w/ | 58.3 (49.9 - 66.5) | 78.8 (75.2 - 82.1) | 77.3 (70.3 - 83.9) |

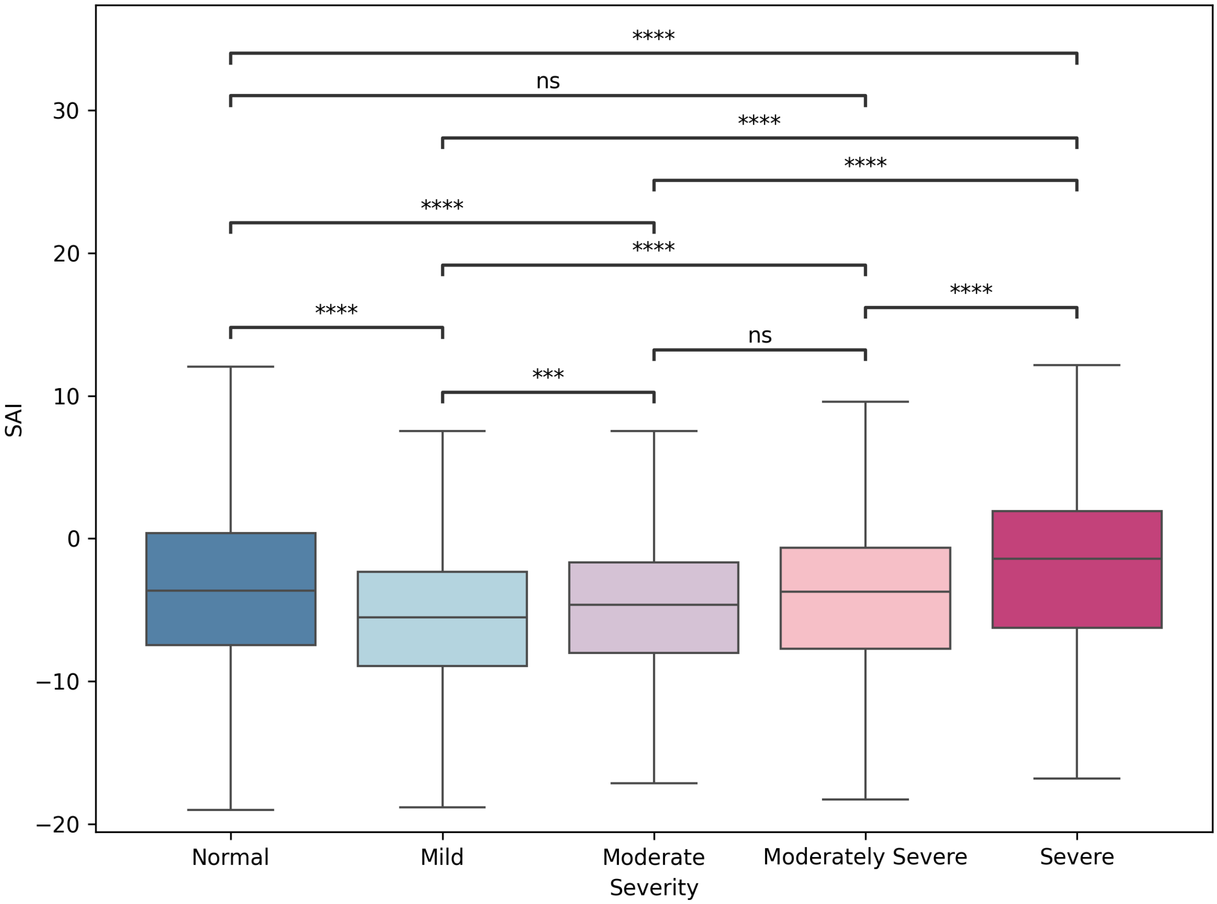


**Figure S5:** Boxplots of SAI compared across the different severity classifications of cognitive impairment, with standard significance reporting ($p < 0.0001$ denoted as ***, $p < 0.001$ denoted as **, $p < 0.05$ denoted as *, and $p < 0.1$ denoted as $\cdot$).

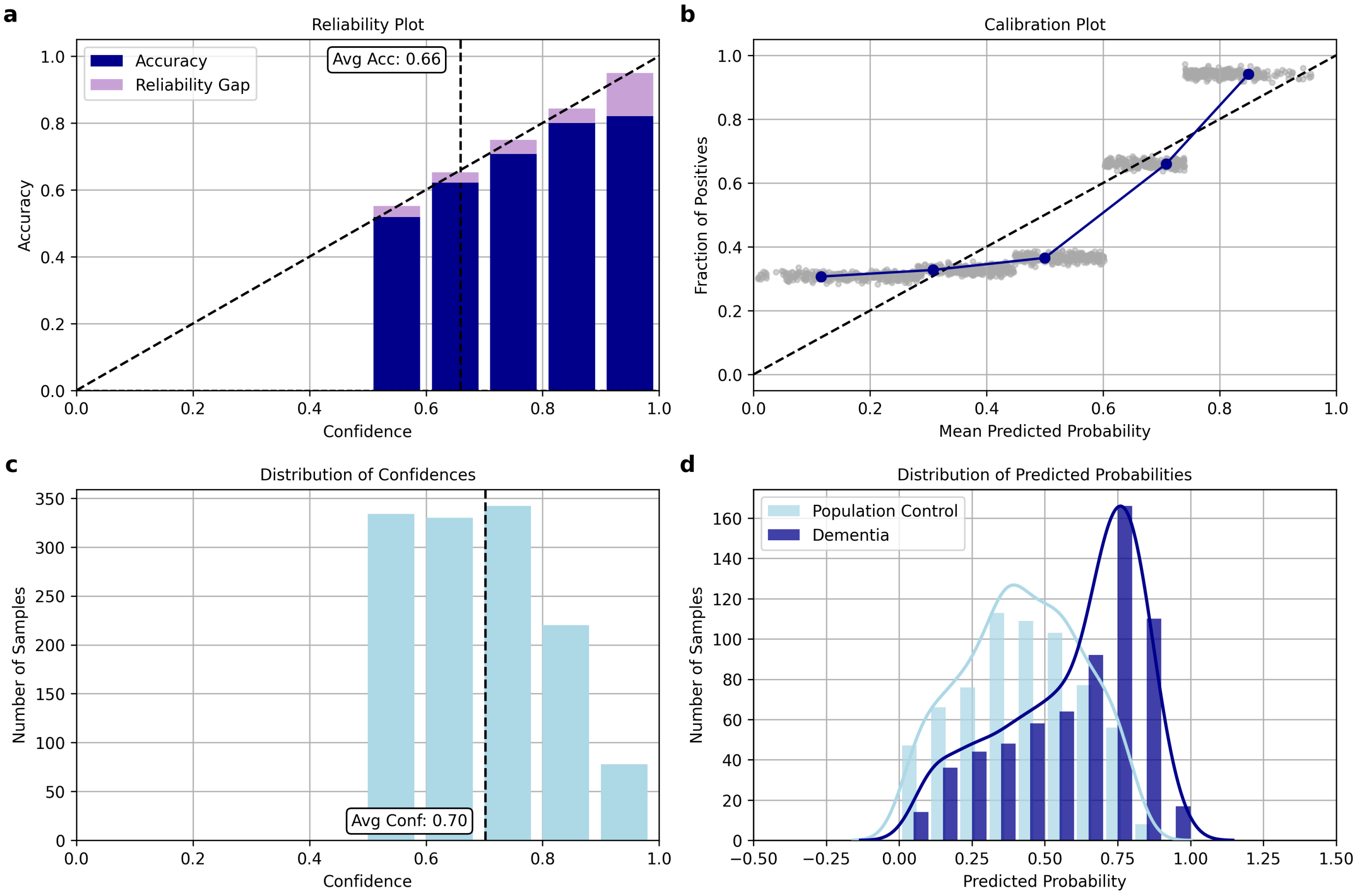


**Figure S6:** Multi-faceted Summary of the Performance of the Dementia Risk Prediction Model. Plot (a) shows the model confidence (on all samples) against accuracy on the held-out test set (n = 1,304, p = 85), with the gaps showing the difference between average accuracy and confidence for a given bin, which would ideally be 0. Plot (c) shows the histogram of confidences reported on the held-out test set. Plot (b) shows the mean predicted dementia risk score (grouped into 5 bins) against the proportion of samples from individuals with dementia, with individual data points depicted in dark grey. Plot (d) shows the distribution of probabilities made by the model for each cohort: Population Control and Dementia.

### S10 Stratification analysis

As first presented by Capstick *et al.* in an analysis of UTI risk,[75] we applied the same stratification approach when reporting our results. We stratified our results into three groups: Green, Amber, and Red, where each group represents low, medium, and high likelihood of a sample coming from an individual in the Dementia cohort, respectively.

We used a 5-fold cross-validation to train the best performing classification model on the training data, deriving dementia risk scores on the validation data so that they can be used to determine the best thresholds for the final model. We then calculate sensitivity, specificity, and precision under different threshold values.

When investigating thresholds, our goal was to ensure that both false negatives and false positives were kept low, so we wanted to maximise sensitivity and specificity, whilst also ensuring that the number of samples stratified into the Green and Red groups was realistic considering prevalence of dementia in older adults in different age ranges, risk of misdiagnosis, and available resources. To achieve this, we varied thresholds with a resolution of 5%, computing these evaluation metrics on the Green and Red groups (negative and positive class groups, respectively) for each pair of thresholds. These thresholds were optimised using the following conditions:

- The maximum difference between the green and amber thresholds must be no more than 50%.
- The range of the green and red groups must be equal.
- The maximum percentage of samples assigned 'Amber' labels must be no more than 50%.
- Finally, to jointly maximise sensitivity and specificity by optimising for the J Youden's statistic:

$$J = (Sensitivity + Specificity) - 1 \tag{7}$$

After employing these restrictions across each split, we found that the best thresholds to apply to our dementia risk scores (with range [0, 100]) were as follows: Green $\epsilon$ [0, 30], Amber $\varepsilon$ [30, 70], and Red $\varepsilon$ [70, 100].

### S11 Performance on female/male splits and across age groups

In this section, we present the model performance on samples from female and male participants and, using the age bands of 50-59, 60-69, 70-79, and 80+ in our dataset. We calculate the accuracy before and after stratification using a binary threshold (where the positive class has a threshold $>$ 50% and the negative class has a threshold of $<=$ 50%) and our chosen thresholds (Green $\epsilon$ [0, 30], Amber $\varepsilon$ [30, 70], and Red $\varepsilon$ [70, 100]) to split the predictions made by the proposed model for male and female participants and for each age group in the held-out test set. For each sex and age group, we also report the likelihood of a positive prediction, as well as the total number and ratio of samples from the Population Control and Dementia cohorts (see Tables S12 and S13).

**Table S12:** Key performance metrics (accuracy, sensitivity, and specificity) on samples from male and female participants in the held-out test set. We report this before and after stratification is performed. We also show the ratio of samples from the Population Cohort (PC) and Dementia (D) cohorts for each sex and the likelihood of a positive prediction.

**Before Stratification**

| Sex | Accuracy | Sensitivity | Specificity | No. of Samples | PC : D | P(Y = 1 — Sex) |
|---|---|---|---|---|---|---|
| **Female** | 72.7 | 93.4 | 50.4 | 494 | 1 : 0.93 | 61.0 |
| **Male** | 61.9 | 53.4 | 69.8 | 810 | 1 : 1.06 | 44.8 |

**After Stratification**

| Sex | Accuracy | Sensitivity | Specificity | No. of Samples | PC : D | P(Y = 1 — Sex) |
|---|---|---|---|---|---|---|
| **Female** | 83.4 | 100 | 42.4 | 229 | 1 : 0.40 | 60.8 |
| **Male** | 65.7 | 58.0 | 73.5 | 443 | 1 : 0.98 | 44.3 |

Applying binary thresholds highlighted a discrepancy in the accuracy according to both the sex and the age group of the participant from which the sample came, with the discrepancy particularly noticeable across age groups due to the difference in

**Table S13:** Key performance metrics (accuracy, sensitivity, and specificity) on samples across the four age bands in the held-out test set. We report this before and after stratification is performed. We also show the ratio of samples from the Population Control (PC) and Dementia (D) cohorts for each age group and the likelihood of a positive prediction.

**Before Stratification**

| Age Group | Accuracy | Sensitivity | Specificity | No. of Samples | PC : D | P(Y = 1 — Age Group) |
|---|---|---|---|---|---|---|
| **50-59** | 80.8 | 0.00 | 81.8 | 245 | 1 : 80.7 | 35.3 |
| **60-69** | 59.3 | 68.6 | 53.6 | 450 | 1 : 1.62 | 51.7 |
| **70-79** | 48.2 | 54.3 | 35.9 | 313 | 1 : 0.49 | 49.2 |
| **80+** | 82.4 | 82.2 | 84.4 | 296 | 1 : 0.12 | 64.6 |

**After Stratification**

| Age Group | Accuracy | Sensitivity | Specificity | No. of Samples | PC : D | P(Y = 1 — Age Group) |
|---|---|---|---|---|---|---|
| **50-59** | 96.3 | 0.00 | 97.5 | 82 | 1 : 81 | 35.0 |
| **60-69** | 71.3 | 77.0 | 64.7 | 251 | 1 : 0.86 | 47.6 |
| **70-79** | 47.6 | 44.9 | 53.3 | 143 | 1 : 0.46 | 60.8 |
| **80+** | 95.1 | 94.8 | 100 | 164 | 1 : 0.07 | 74.4 |

the number of samples from the Dementia and Population Control cohorts for each age group. We see that female participants and those age 70 years and older are more likely to come from the Dementia cohort than male participants, while 50 to 69 year olds are more likely to come from the Population Control cohort. This difference in label proportions makes a disparity in metrics on different demographics inevitable. After stratification, we see that accuracy has increased for each sex and all but one age group (70-80), while the discrepancy is seen to increase between older and younger age groups.

This experiment also highlighted both the advantages and risks of tuning dementia risk score thresholds on individual sexes and age groups based on a chosen fairness metric for a specific research scenario or setting. For example, Tables S12 and and S13 show the likelihood of a positive prediction (Y = 1) by sex (P(Y = 1 | Sex)) and age group ($P(\hat{Y} = 1$ | Age Group)) using our proposed model with binary thresholds and chosen thresholds. This is referred to as demographic parity, and we see here that while our model has moderate demographic disparity across samples from male and female participants, before and after stratification, and also across age groups, increasing more so in older age groups.

While stratification does not improve statistical parity, we see that accuracy does increase across both sexes and three of the four age bands, making the stratification beneficial overall, although the trade-off in fairness should be considered and ideally both accuracy and fairness optimised for optimal clinical usability.

### S12 Effect of seasonality

To assess potential seasonal bias in our model outputs, we conducted ANOVA on both our age estimations and dementia risk predictions. While no statistically significant differences were observed across seasons in the SAI of the Pilot High-risk cohort ((F(3, 466) = 1.46, p = 0.23)), they were observed in the Population Control (F(3, 3,087) = 14.0, p = 0.00) and Dementia (F(3, 3,136) = 6.73, p = 0.00) cohorts, with follow-up pairwise comparisons using Tukey's HSD indicating that the observed seasonal variation in the Population Control cohort was driven by a difference between the Winter and Summer (p = 0.00) and Summer and Autumn (p = 0.00) and in the Dementia cohort by a difference between Winter and Summer ($p < 0.01$) and Winter and Autumn ($p < 0.001$). However, this effect was not present in the risk scores within the combined (Population Control and Dementia) cohort (F(3, 1,300) = 0.80, p = 0.49) or the Pilot High-risk cohort (F(3, 466) = 0.56, p = 0.57).
